# Efficient and accurate mixed model association tool for single-cell eQTL analysis

**DOI:** 10.1101/2024.05.15.24307317

**Authors:** Wei Zhou, Anna S.E. Cuomo, Angli Xue, Masahiro Kanai, Grant Chau, Chirag Krishna, Ramnik J. Xavier, Daniel G. MacArthur, Joseph E. Powell, Mark J. Daly, Benjamin M. Neale

**Affiliations:** Psychiatric and Neurodevelopmental Genetics Unit, Center for Genomic Medicine, Massachusetts General Hospital, Boston, MA, USA; Stanley Center for Psychiatric Research, Broad Institute of MIT and Harvard, Cambridge, MA, USA; Program in Medical and Population Genetics, Broad Institute of MIT and Harvard, Cambridge, MA, USA; Institute for Molecular Medicine Finland, University of Helsinki, Helsinki, Finland; Novo Nordisk Foundation Center for Genomic Mechanisms of Disease, Broad Institute of MIT and Harvard, Cambridge, MA 02142, USA; Translational Genomics Program, Garvan Institute of Medical Research, Sydney, NSW, Australia; Centre for Population Genomics, Garvan Institute of Medical Research and UNSW Sydney, Sydney, New South Wales, Australia; Centre for Population Genomics, Murdoch Children’s Research Institute, Melbourne, Victoria, Australia; School of Biomedical Sciences, University of New South Wales, Sydney, NSW, Australia; Infectious Disease and Microbiome Program, Broad Institute of MIT and Harvard, Cambridge, MA, USA; Department of Molecular Biology, Massachusetts General Hospital, Boston, MA, USA; Center for Computational and Integrative Biology, Massachusetts General Hospital, Harvard Medical School, Boston, MA, USA; Analytic and Translational Genetics Unit, Department of Medicine, Massachusetts General Hospital, Boston, MA, USA; UNSW Cellular Genomics Futures Institute, University of New South Wales, Sydney, NSW, Australia

## Abstract

Understanding the genetic basis of gene expression can help us understand the molecular underpinnings of human traits and disease. Expression quantitative trait locus (eQTL) mapping can help in studying this relationship but have been shown to be very cell-type specific, motivating the use of single-cell RNA sequencing and single-cell eQTLs to obtain a more granular view of genetic regulation. Current methods for single-cell eQTL mapping either rely on the “pseudobulk” approach and traditional pipelines for bulk transcriptomics or do not scale well to large datasets. Here, we propose SAIGE-QTL, a robust and scalable tool that can directly map eQTLs using single-cell profiles without needing aggregation at the pseudobulk level. Additionally, SAIGE-QTL allows for testing the effects of less frequent/rare genetic variation through set-based tests, which is traditionally excluded from eQTL mapping studies. We evaluate the performance of SAIGE-QTL on both real and simulated data and demonstrate the improved power for eQTL mapping over existing pipelines.

## INTRODUCTION

Transcriptomes have been extensively measured by RNA sequencing (RNA-seq) followed by expression quantitative trait locus (eQTL) mapping across multiple cell types and tissues to identify genetic variants associated with gene expression^1–3^. eQTL analysis provides evidence on functional genetic variants and facilitates understanding the molecular basis for human diseases when integrated with genome-wide association studies (GWASs)^4^. More recently, single-cell RNA-seq (scRNA-seq) methods have enabled a more fine-grained view of cellular diversity, and single-cell eQTL mapping can help uncover gene regulation at a deeper resolution of cell types^5^. Indeed, when studying the effects of genetic variation on expression using bulk transcriptomes across multiple cell types, it is virtually impossible to differentiate between true transcriptomic effects as opposed to effects on cell type proportions^6^. Further, bulk modelling fails to consider intra-individual cell-to-cell variability.

The first generation of single-cell eQTLs considered aggregated (typically, mean) gene expression from multiple cells per individual, using a so-called “pseudobulk” approach. This approach has provided important insights into the genetic basis of cell type-specific gene expression across several tissues^7–14^, but has limitations. In particular, the pseudobulk approach does not appropriately model the intra-individual cell-to-cell variability. Thus, it cannot leverage information shared across cells with similar profiles and lose power.

More recently, new studies have proposed approaches that model the expression levels of single cells directly^15–17^ and effectively account for key characteristics of these data; first, they can handle the presence of multiple cells from the same individual using a random effect term, and second, they appropriately model the non-normally distributed single-cell counts by adopting discrete count distributions (*e*.*g*., Poisson). Yet, they still have several limitations. Most notably, these methods are computationally inefficient. They are typically used only for downstream interpretation of cell state or cell transition specific effects, following faster pseudobulk methods which are still employed for the main analysis^15,16^. Additionally, the increasingly larger number of cells in current studies^7,17,18^, which is fast approaching multiple millions as new datasets are generated^17^, poses more challenges in computational efficiency. Efficient memory usage and speed are particularly relevant for single-cell eQTL studies, which are only in their infancy and where the number of cells and individuals is ever-increasing. Moreover, thousands of phenotypes (=genes) are considered in eQTL analyses, and tens of cell types, further increasing the number of tests that need to be performed. Current methods are not scalable at the genome-wide scale thus cannot be used to identify *trans*-eQTLs, even though previous studies suggest *trans*-eQTLs explain a large proportion of gene expression variability^19–21^. Finally, bulk and single-cell eQTL studies have historically been limited to common genetic variants (population minor allele frequency; MAF > 5%). This is largely due to data availability, with most studies relying on genotype arrays and imputation, which is limited to selected tagged (common) loci on the genome. Generally, small sample sizes also limit the exploration of rare variations. From a methodological perspective, eQTL methods typically perform single-variant tests (where only one variant is tested at a time), which are underpowered for low frequency variants. As the number of studies employing whole genome sequencing grows with increased sample sizes, opportunities arise to explore the role of rare genetic variations on gene expression. However, this also highlights the necessity for developing more efficient methodologies.

Here, we propose SAIGE-QTL, an efficient and scalable approach for single-cell QTL mapping, that addresses the above mentioned limitations. SAIGE-QTL builds on SAIGE^22^, SAIGE-GENE^23^, and SAIGE-GENE+^24^, a previously proposed suite of scalable and efficient generalised linear mixed model tools, already addressing similar challenges in the context of GWAS analyses. First, SAIGE-QTL implements a mixed-effect model, thus modelling the repeated sample structure resulting from multiple cells derived from the same individual, as well as the relatedness of the individuals considered, if any, via the inclusion of a genetic relatedness matrix (GRM) in the model. Second, it uses a Poisson mixed model, effectively modelling the single-cell expression profiles as discrete counts. Third, SAIGE-QTL is fast and scalable for large datasets and can efficiently test thousands of genes, tens to hundreds of cell types, and millions of cells and genetic variants. Importantly, this allows for *trans*-eQTL mapping, where we test for the effects of variants that are distant or even on a different chromosome than the target gene as well as *cis*-eQTL mapping, where the variants tested are in close vicinity of the target gene. Finally, SAIGE-QTL can test for the effects of rare and less frequent genetic variants (MAF ≤5%) on gene expression through the implementation of efficient set-based tests and employs the saddle-point approximation (SPA^25^) for better controlled type I error rates. Altogether, we introduce a novel method for single-cell eQTL mapping that overcomes several limitations of existing approaches, and demonstrate its utility on both simulated and real data, using the largest available population-scale single-cell dataset, the OneK1K cohort^7^, showing notable improvements in performance compared to existing tools.

## RESULTS

### Overview of Methods

The SAIGE-QTL method contains three main steps for each gene (**Supplementary Figure 1, Supplementary Note**): (1) fitting the null Poisson mixed model, which includes individual-level covariates, such as sex, age, and ancestry PCs for donors, and cell-specific covariates, such as expression-based PEER^26^ factors, as fixed effects and accounts for intra-individual cell-to-cell variability using a random effect; (2) a. testing for the association between each common genetic variant (*e*.*g*., MAF > 5%) and gene expression using score tests; b. conducting set-based tests for less frequent/rare variants with *e*.*g*., MAF ≤5%, including Burden tests, SKAT^27^, and ACAT-V^28^, each having superior power in various scenarios of effect directions and sparsity levels followed by an ACAT-O test^28^ to combine p-values. (3) calculating a gene-level p-value for each *cis*-eQTL region using the ACAT-V test^28^, which combines variant-level p-values (from Step 2a) using a Cauchy combination that is robust to the correlation among individual p-values and the sparsity of effects.

Step 1 iteratively estimates the model parameters using the penalised quasi-likelihood (PQL^29^) method and the computationally efficient average information restricted maximum likelihood (AI-REML) algorithm^30^, also used in SAIGE^22^ and GMMAT^31^. Several optimisation strategies have been applied to make fitting the null Poisson mixed model practical for large single-cell RNA-seq data sets with millions of cells across thousands of individuals, such as OneK1K^7^ (**Methods; Supplementary Note**). For example, the preconditioned conjugate gradient (PCG^32^) approach for solving linear systems is used to avoid inverting the *N* × *N* matrices through the matrix decomposition, which cost *O*(*N*^3^), where *N* is the total number of cells in scRNA-seq studies and can range from hundreds of thousands to millions, increasing with study sizes. The computation time is about *O*(*nNP*) times the number of iterations for PCG to converge, where *n* is the number of individuals (donors) and *P* is the number of covariates including the intercept. The *n* × *N* cell by individual matrix indicating which individual each cell belongs to is stored in memory as a sparse matrix with 1s. Once the null model is fitted for a gene in a cell type, it can be used to test the association of any genetic markers or regions for that gene’s expression in that cell type.

In Step 2, for common variants, the score test is performed based on model residuals and parameters estimated in Step 1 to test the association between each genetic variant and the expression level of the gene of interest. The overall cost of computing the variance of the score statistic for all variants is extremely high since the inversion of a *N* × *N* matrix is needed for each variant. To overcome this issue, SAIGE-QTL uses the variance ratio approximation derived under the Poisson mixed model by extending that used in SAIGE and other GWAS methods^33–35^. Two variance ratios are computed using a random subset of genetic markers, which have been previously shown to be approximately constant for variants with MAC ≥ 20 for linear mixed models^33,34^ and generalised linear mixed models^22^ (**Methods** and **Supplementary Note**). These include a ratio of the full variance of the score statistic to the variance without any random effects, as well as a ratio of the full variance of the score statistic to the variance with random effects (which is cheaper to compute than the full variance), Notably, estimates of the second ratio have lower variation compared to the first, as previously demonstrated in the SAIGE-GENE method^23^. To improve the computation efficiency while maintaining test accuracy, after fitting the null Poisson mixed model, SAIGE-QTL first approximates the full variance of score statistics using the variance calculated without random effects and the ratio of the two, similar to previous mixed model methods for GWASs, *e*.*g*., BOLT-LMM^33^, GRAMMAR-Gamma^34^, and SAIGE^22^. If the association p-value is lower than a cutoff, *e*.*g*., 0.05, SAIGE-QTL then estimates the full variance of score statistics using the variance calculated with random effects and the ratio of the two and re-calculates the p-value. In addition, SAIGE-QTL approximates the distribution of the score test statistics using SPA to obtain more accurate p-values than using the Normal distribution.

For less frequent/rare variants with MAF ≤5%, after fitting the null model in Step 1, set-based association tests, including Burden, SKAT^27^, and ACAT-V^28^ tests, are performed in Step 2b (**Supplementary Figure 1**). SAIGE-QTL improves the power^23,24^ by allowing for incorporating multiple MAF cutoffs, functional annotations, and weights, *e*.*g*., based on the distance to the transcription start site (TSS), similar to approaches suggested by others^36,23,24^. Results from different tests are combined using the minimum p-value or Cauchy combination method^28^ as used in SAIGE-GENE+^23,24^.

In Step 3, a gene-level p-value is estimated based on the Cauchy combination of variant-level p-values. This approach was first suggested by the APEX^36^ authors and provides highly concordant gene-level p-values compared to those obtained using the Beta approximation method described in the FastQTL paper^37^ and implemented in TensorQTL^38^ (Pearson’s R^2^ =1 when tested on p-values from TensorQTL applied to OneK1K, see below and **Supplementary Figure 2**).

#### Computation and memory cost

We evaluated the computation cost of SAIGE-QTL when mapping eQTLs from the largest-to-date cohort-scale scRNA-seq dataset, OneK1K^7^. Briefly, the OneK1K dataset consists of scRNA-seq from over 1.2 million immune cells of 982 unique individuals, categorised into 14 distinct (sub)cell types and matched genotypes (**Methods**). In total, 16,250 genes are expressed in at least 10% of the donors in at least one cell type (range: 6,326-15,400 across different cell types) (**Supplementary Table 1**), and 12,108,282 single-base variants are called or imputed across the frequency spectrum (5,328,917 with MAF>=5%, 6,779,365 with MAF<5%) for all autosomal chromosomes (1-22).

The cost of Step 1 (fitting the null model) ranged between 34 and 1,569 CPU hours and 0.37 and 1.96Gb of memory usage across different cell types, with the differences being largely driven by the number of cells present for different cell types. The projected overall computation time (**Supplementary Table 2**) for Step 2 to test eQTLs in *cis*, considering a window of +/− 1M base pairs around the gene body, on all genetic variants with MAF > 5% for 20,000 genes was between 28 and 40 CPU hours across the 14 cell types. The estimated time was projected for testing 20,000 genes in each cell type based on the actual time cost on genes expressing in at least 10% of the donors (**Supplementary Table 1**). We benchmarked the *cis*-eQTL mapping for gene *RPL23A* in naïve B cells (“B_IN_”)) as an example to compare SAIGE-QTL to the glmer function in the lme4 R package based on Poisson mixed models^39^, which was used previously by Nathan et al.^15^. SAIGE-QTL reduced the computation time by 59 times (2mins 50s for Step 1 and Step 2 by SAIGE-QTL vs. 2.83 hrs by glmer for testing 2,019 markers with MAF > 5%).

We also evaluated the time cost when testing for effects of all common variation genome-wide for three key representative cell types from the OneK1K dataset: CD4 naive and central memory T cells (“CD4_NC_”) as the most abundant cell type, with 463,528 cells in total; immature and naïve B cells (“B_IN_”) as an intermediate cell type (82,068 cells) and Plasma, as the least abundant cell type from PBMCs in the OneK1K dataset (3,625 cells). Testing 5.3 million variants across the genome with MAF > 5% for the 20,000 expressed genes was projected to take 630, 719 and 1013 CPU hours for Plasma, B_IN_, and CD4_NC_ respectively (**Supplementary Table 2**). As is shown, in genome-wide eQTL mapping, each genetic marker can be tested for multiple genes (phenotypes) in each Step 2 job. Reduction in the overhead of genotype reading substantially saves computation time. In addition, parallel computation has been implemented to reduce computation time further when using multiple CPUs for Step 2 jobs that test multiple genes.

#### Type I error rates/False positive calibration

To evaluate the false positive calibration of SAIGE-QTL, we estimated the type I error rates of SAIGE-QTL for mapping eQTLs using single-cell RNA-seq data by following the steps described in the “**Data simulation**” subsection of the **Methods** section using the real single-cell read counts of gene expression and permuted genotypes in the OneK1K data.

To evaluate the calibration of SAIGE-QTL across cell type abundances, gene expression levels (measured in terms of sparsity, defined as the proportion of zero read counts), we considered the three cell types described above and randomly selected 50 genes with low (< 20%), medium (45 ~ 55%) and high (> 80%) sparsity levels in each cell type (**Supplementary Table 3**). For each gene, we first fitted a null Poisson mixed model using the real read counts for the gene from cells across individuals in Step 1 and then permuted the individual IDs to conduct the single-variant association tests on 405,663 genetic variants on chromosome 1 with MAF > 5% in Step 2a. In this way, we disrupted real association signals while retaining the per-individual read counts distribution across cells for the gene expression. We repeated the permutation 250 times and estimated the empirical type I error rates as the proportion of p-values smaller than the given *α* level based on 1.01×10^9^ tests for each gene. Empirical type I error rates were estimated with 50 genes combined on each sparsity level for each cell type at 7 different *α* levels from 5×10^−8^ to 0.05 (**Supplementary Table 4**). Together with plots of the genomic control lambda values corresponding to the 1st and 50th percentile (median) in **Supplementary Figure 3A** and the quantile-quantile (Q-Q) plots for 9 random genes in **Supplementary Figure 3B**, our results suggested that SAIGE-QTL has well-controlled type I error rates for the single-variant association tests, with slightly deflated type I error rates when the sparsity level is medium to high and in less abundant cell types (Plasma and immature and naïve B cells).

To evaluate type I error rates of the ACAT-V tests in Step 3 that are used in SAIGE-QTL to obtain a p-value for each gene at the *cis*-region, we then performed the ACAT-V test to combine single-variant p-values for variants with MAF > 5% in each gene region (a window of +/− 1M base pairs around the gene body) on chromosome 1 with permuted individual IDs. We repeated the tests for 250 times with permutation of individual IDs in Step 2a for three randomly selected genes from each sparsity group and cell type. As shown in **Supplementary Table 5** and **Supplementary Figure 4**, similar to single-variant tests, type I error rates were well controlled for the gene-level ACAT-V tests with a slight deflation.

We also performed the set-based tests for rare genetic variants with MAF ≤5% in each gene region on chromosome 1 in Step 2b. The tests were repeated for 250 times with permuted individual IDs and we observed well calibrated set-based tests in SAIGE-QTL for genes from different sparsity groups across three cell types as presented in **Supplementary Figure 5**.

### Association analysis of OneK1K scRNA-seq and imputed genotypes

To demonstrate the application of SAIGE-QTL to real-world scRNA-seq cohorts, we applied it to data from the OneK1K project^7^, which includes scRNA-seq from over 1.2 million immune cells of 982 unique individuals, categorised into 14 distinct (sub)cell types, with matched genotypes. We mapped *cis* eQTLs (considering all SNPs of minor allele frequency >5% and within a +/− 1Mb window around the gene body) for all genes expressed in at least 10% of the donors (in the relevant cell type) (**Supplementary Table 1**).

We used the Poisson mixed model from SAIGE-QTL, modelling single-cell counts directly, as opposed to aggregating counts at the donor level using a “pseudobulk” approach, as conducted in the original paper^7^ (using Matrix eQTL^40^; **Methods**). We used the same covariates as in the original model, correcting for sex, age, 6 genotype PCs and 2 cell type-specific expression PEER^26^ factors from the original publication. We observed well-calibrated p-values in SAIGE-QTL, with genomic control lambda values around one in the three representative cell types (**Figure 1A**). Across all 14 cell types, SAIGE-QTL identified 17,218 eGenes across all cell types (FDR<5%, as described below and in Methods), representing 5,894 unique genes of which 2,447 (42%) were specific to one cell type only (**Supplementary Figure 6**).

**Fig. 1:**
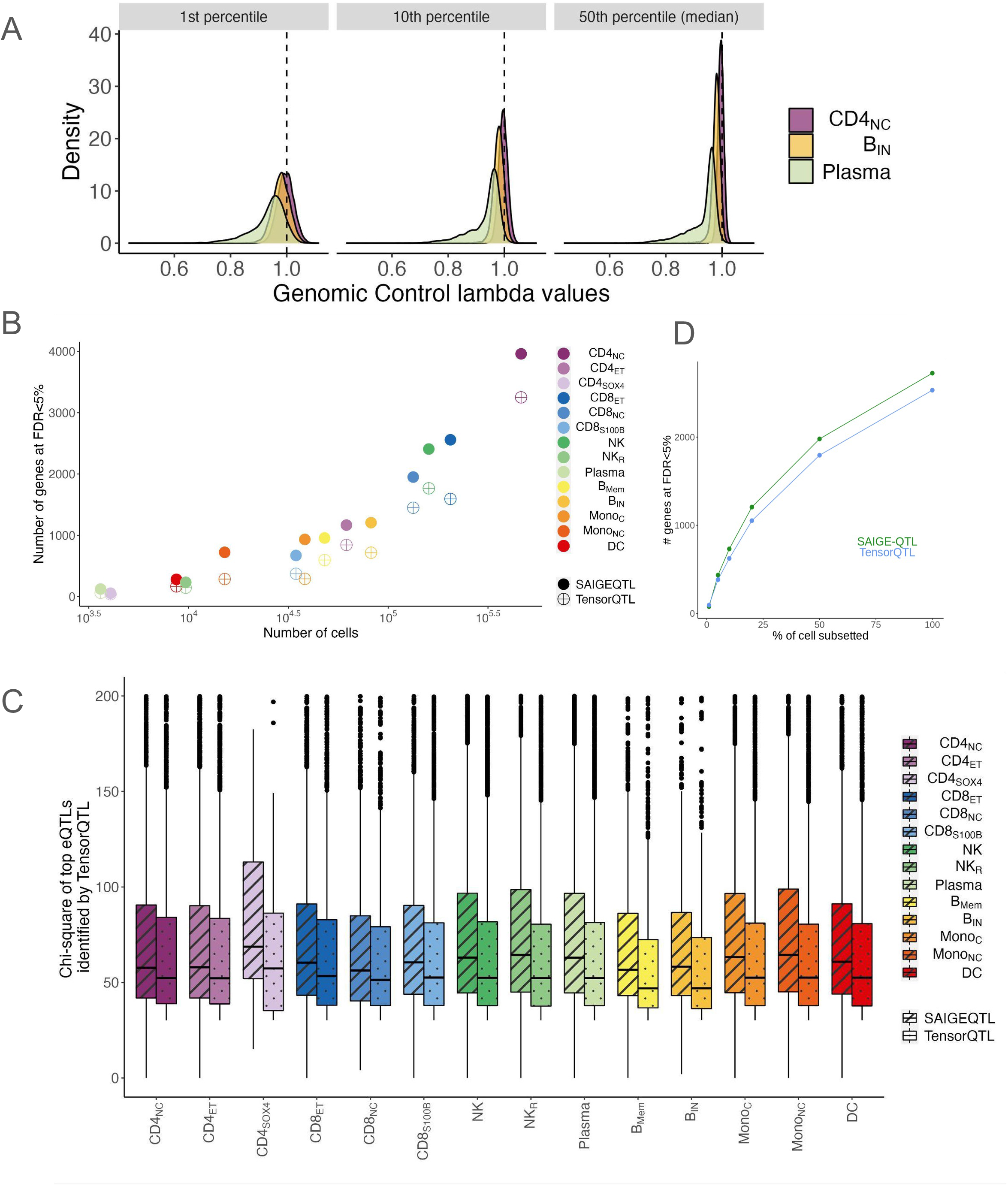
Performance evaluation of SAIGE-QTL. A. Calibration (genomic control lambda values) across three representative cell types. B. Scatter plot of power (number of eGenes identified at FDR<5%, y axis) and number of cells (x axis) between SAIGE-QTL (solid dots) and TensorQTL (crosses) across 14 cell types. C. For each of the 14 cell types, boxplots of chi squared statistics for the top eQTL in SAIGE-QTL (diagonal lines) and TensorQTL (dots). When the top variant was different between methods, the TensorQTL top variant was selected. y axis is truncated at the maximum value 200. D. Scatter plot of power (number of eGenes identified at FDR<5%, y axis) and percentage of cells subsampled for the most abundant cell type (CD4_NC_, **Methods**).

#### SAIGE-QTL identified more eGenes than the pseudobulk approach

We compared SAIGE-QTL with the widely used tool TensorQTL^38^, which we ran using (mean) pseudobulk counts and the same covariates and window size as above across all 14 cell types (**Methods**). We obtained good concordance of the nominal p-values between the two methods (Pearson’s correlation ranges from 0.9 to 0.97, **Supplementary Figure 7A**). We also observed that TensorQTL provides more significant nominal p-values than SAIGE-QTL for eQTLs with extremely significant p-values, *i*.*e*., p-values < 10^−100^, while SAIGE-QTL tends to have more significant p-values than TensorQTL for those less significant eQTL signals as shown in **Supplementary Figure 7B**. Among genes with similar sparsity levels across cells, those with a higher sparsity level after pseudobulking tend to have more significant nominal association p-values in TensorQTL than SAIGE-QTL (**Supplementary Figure 8**). Effect size estimates were concordant too, though to a lesser extent, which can be explained by the different underlying models with different units (Gaussian for pseudobulk vs Poisson model for read counts from individual cells; Pearson’s correlation ranges from 0.35 to 0.71, **Supplementary Figure 9**).

To obtain gene-level p-values for TensorQTL, we used the Beta approximation method described in the FastQTL paper^37^, using 10,000 permutations as recommended by the authors (**Methods**). For SAIGE-QTL, we used the ACAT-V test to combine correlated p-values using the Cauchy combination^28,41^, as described above. In both cases, we further corrected for multiple testing across all genes tested (for each cell type) using the q-value method^42^, reporting results at FDR<5%. For both sets of results, we confirmed a clear link between the number of cells available for each cell type and the number of eQTLs detected (Pearson’s correlation = 0.96, and 0.97 for SAIGE-QTL and TensorQTL, respectively; **Figure 1B**), with SAIGE-QTL detecting 48.8% more eGenes (*i*.*e*., genes with at least one eQTL) compared to TensorQTL (17,218 vs 11,569 eGenes across all 14 cell types) (**Supplementary Table 6A**). Improved power of SAIGE-QTL compared to TensorQTL was confirmed by comparing the chi-square statistics of the most significant genetic variants with nominal p-value < 5×10^−8^ in eGenes across the two methods, which ranged between 3.8% and 19.2% higher for SAIGE-QTL across all 14 cell types (**Figure 1C**). If the two methods identified different top variants, the ones identified by TensorQTL were included in the comparison. As the ACAT-V test can provide slightly more significant p-values than Beta permutation (**Supplementary Figure 2A**), we also applied ACAT-V on nominal p-values from TensorQTL (**Supplementary Figure 2B**). We observed that TensorQTL with ACAT-V detected 10.4% more eGenes than with Beta permutation (12,777 vs 11,569) (**Supplementary Table 6B**). Other analyses in this paper used results from TensorQTL with Beta permutation, which the original TensorQTL program used.

To further evaluate the power of our method, we assessed the effect of the numbers of cells available for association. Considering the most abundant OneK1K cell type, CD4_NC_, we considered different levels of down-sampling, considering subsets of 1, 5, 10, 20 and 50% of all cells (keeping the cell-to-donor ratio intact; **Methods**). In all cases, we ran SAIGE-QTL after selecting all genes expressed in at least 1% of all cells considered (post-subsetting). Additionally, we generated pseudobulk counts (by considering the mean expression level across cells for each individual, gene and cell type) for each of the 5 cell subsets and also ran TensorQTL for the same genes. As expected, the number of eGenes detected decreased as the number of cells considered decreased, and SAIGE-QTL consistently identified more significant eGenes than TensorQTL (75% more on average, **Figure 1D**).

Reassuringly, we retained a large overlap between the sets of eGenes (at FDR<5%) identified across the two methods, with on average 94% of eGenes identified by TensorQTL found by SAIGE-QTL also (across 14 cell types, range: 87-96%, **Figure 2A**). As an example of an eQTL found using SAIGE-QTL and missed by TensorQTL, we identified an eQTL (rs1375493) for the inflammatory bowel disease (IBD)-associated *ITGA4* locus, which is a well known regulatory signal, and is able to recapitulate the monocyte-specificity of the eQTL, which has been previously shown^4^ (**Figure 2B** and **Supplementary Figure 10**). Similar to what is done in the original OneK1K study, we next performed five rounds of conditional association analysis (conditioning on previously significant variants to identify additional independent signals), which identified 4,647 additional eQTLs across the 14 cell types in OneK1K (for 2,435 genes, **Supplementary Figure 11**).

**Fig. 2:**
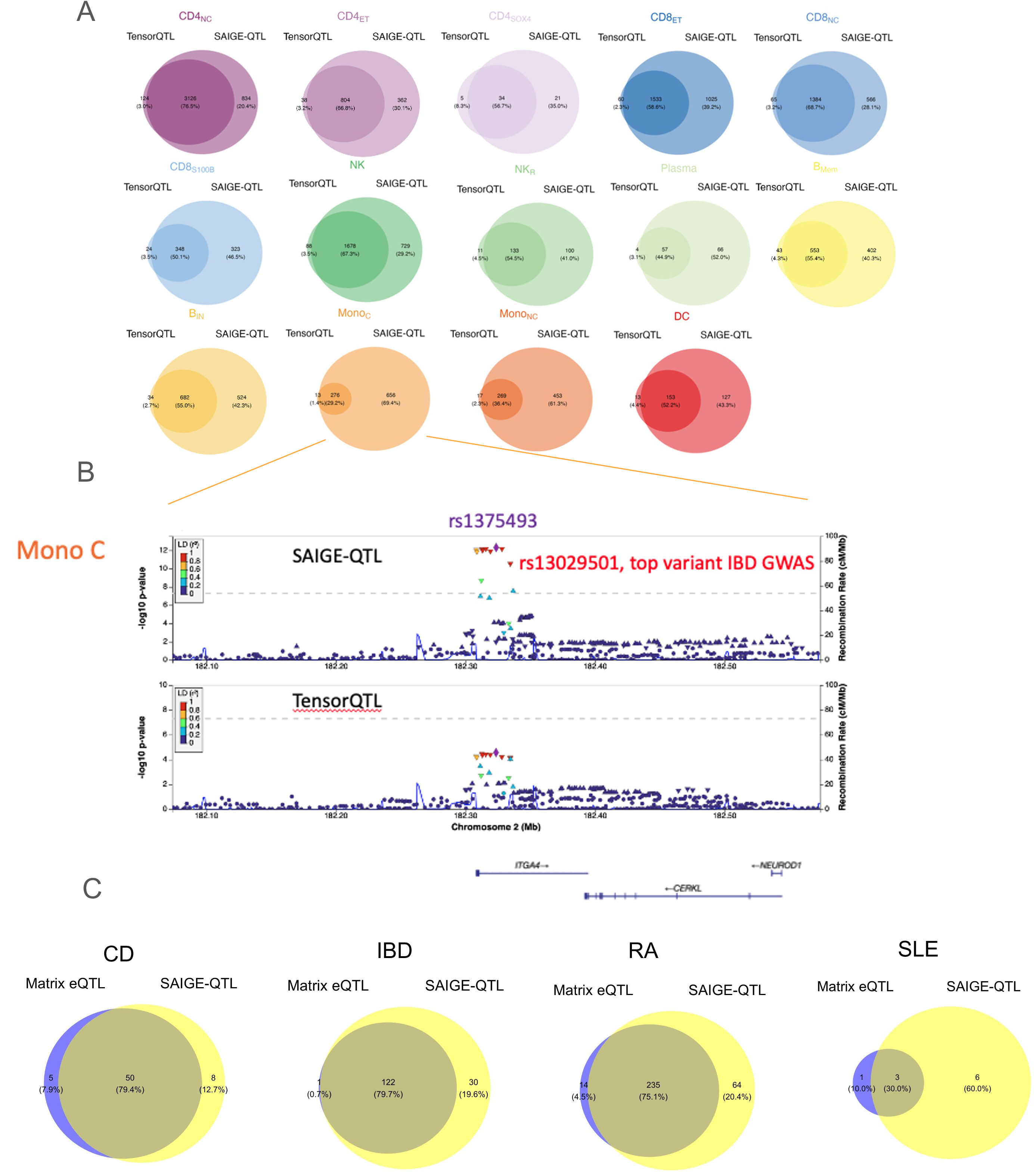
SAIGE-QTL single-cell eQTL mapping identifies more eQTLs than pseudobulk methods. A. Venn diagrams representing the overlap of significant eGenes (FDR<5%) identified by SAIGE-QTL and TensorQTL across all 14 cell types. B. Example of disease-specific eQTL detected by SAIGE-QTL and not by TensorQTL (*ITGA4* locus, monocytes-specific eQTL (see **Suppl. Fig. 10**)). C. Among eGenes identified by SAIGE-QTL and TensorQTL across 14 cell types, gene-cell type pairs that are identified by the Summary-data–based Mendelian randomization (SMR) framework to be associated with four autoimmune disease risks. CD: Crohn’s disease, IBD: inflammatory bowel disease, RA: rheumatoid arthritis, SLE: systemic lupus erythematosus.

#### SAIGE-QTL identified additional immune disease-associated loci mediated by gene expression

We then use a Mendelian randomization approach to identify loci contributing to the autoimmune disease through gene expression changes in any of the 14 cell types. The SMR tool (**Methods**) was used to conduct the analysis using eQTL results by the SAIGE-QTL and the original Matrix eQTL^40^ and GWAS summary statistics for four autoimmune diseases, Crohn’s disease (CD)^43^, inflammatory bowel disease (IBD)^43^, rheumatoid arthritis (RA)^44^, and systemic lupus erythematosus (SLE)^45^. We found that the SMR p-values are very concordant when using results from the two methods, with results from SAIGE-QTL identifying more SMR associations compared to Matrix QTL across all four diseases (n=58 vs n=55 for CD, 152 vs 123 for IBD, 299 vs 249 for RA, and 9 vs 4 for SLE. **Supplementary Table 7**), with on average 90% gene-cell type combinations using eQTL results from Matrix eQTL also identified when using eQTL results from SAIGE-QTL (**Figure 2C**). In addition, using results from SAIGE-QTL, more unique loci were detected to contribute to the disease of interest through gene expression change in one or more cell types (n=17 vs n=14 for CD, 34 vs 28 for IBD, 77 vs 63 for RA, and 5 vs 4 for SLE, **Supplementary Figure 12)**. Notably, several loci identified using eQTL results from SAIGE-QTL but missed by using results from Matrix eQTL act through changes in expression of genes that are known to be implicated in the autoimmune diseases. For example, the locus at 9q21 was shown to affect the risk of CD and IBD through changes in *CARD9* expression in monocytes (Mono_C_), where previous studies suggested the dysfunction of *CARD9* may contribute to the pathogenesis of IBD^46,47,48.^ The locus 20q13 was identified to causally contribute to RA through the gene expression change of *CD40* in plasma and the *CD40* pathway has been long implicated in different studies for RA^49–51^, which is a promising treatment target^52–54^.

#### SAIGE-QTL enables genome-wide scan for identifying Trans-eQTLs

Scalable and fast, SAIGE-QTL enables genome-wide scans of genetic effects on single-cell expression, effectively allowing testing for *trans* eQTLs (where the genetic variant tested is far away or even on a different chromosome compared to the target gene). We mapped *trans* eQTLs for the three representative cell types selected above (CD4_NC_, B_IN_, Plasma). In total, we identify 413 *trans* eQTLs for 390 genes across the three cell types (at p<5e-8, MAF ≥10%, **Figure 3A**, results for p<5e-6 reported in **Methods** and **Supplementary Figure 13**). As an example, rs3924376 on chromosome 16 is a *cis* eQTL in CD4_NC_ for *SPNS1*, a gene involved in mitochondrial homeostasis^55^, and a *trans* eQTL for *MRPL32* a chromosome 7 gene encoding Mitochondrial ribosomal protein L32 (**Figure 3B**).

**Fig. 3:**
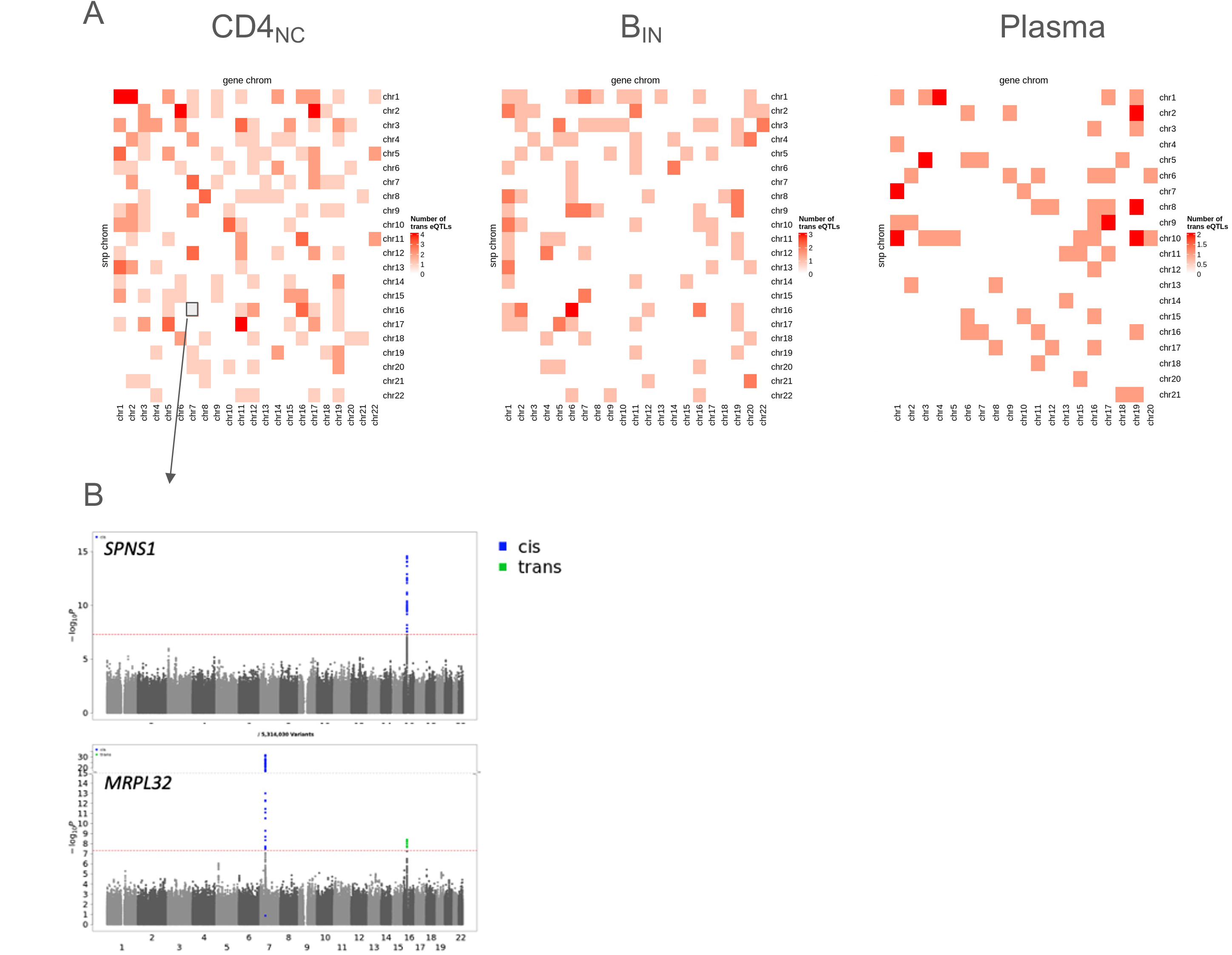
Computational efficiency of SAIGE-QTL allows genome-wide scans to identify *trans* eQTLs. A. Overview of trans effects (heatmap of chromosome by chromosome relationships) in 3 representative cell types (p<5e-8). B. Example(s) of a variant that has a cis eQTL effect (blue) on *SPNS1* (top) and also a trans eQTL effect (green) on a different gene (*MRPL32*, bottom) in, CD4_NC_. Shown are Manhattan plots for each of the two genes.

#### Rare-variant eQTL signals identified by SAIGE-QTL

Finally, we tested for the effects of rare variants on single-cell expression across all 14 cell types. In total, 6,779,365 rare SNPs (MAF ≤5%) could be called from the SNP array data from the OneK1K paper^7^. For each gene expressed in at least 10% of donors for a given cell type, we considered all rare variants within a *cis* window up and downstream of the gene body (+/−1Mb). We performed set-based tests (considering all variants in the window at once), including Burden, SKAT and ACAT-V. Overall, we identified 5,541 rare (MAF≤5%) variant eGenes (RV-eGenes) across all 14 cell types (2,317 unique genes, **Figure 4A, Supplementary Figure 14**). Of these, 483 (21%) were independent of common eQTL signals from the same genes, which remained significant (FDR < 0.05) after conditioning on all independent significant common eQTLs (**Supplementary Figure 15**). We also ran the set-based tests using weights based on the distance from the transcription start site (“dTSS”, **Methods**), finding on average 68% (range: 50-89% across cell types) more significant effect when compared to using equal weights, and 77% (50-112%) when compared to the traditional weighting of variants following a Beta(1,25) distribution (**Figure 4B**). Compared to the common variants, we observe about a third of the number of eGenes identified (mean=0.33, min=0.26, max=0.49) across different cell types, with a very strong correlation, due to the number of cells available for each cell type (**Figure 4C**).

**Fig. 4:**
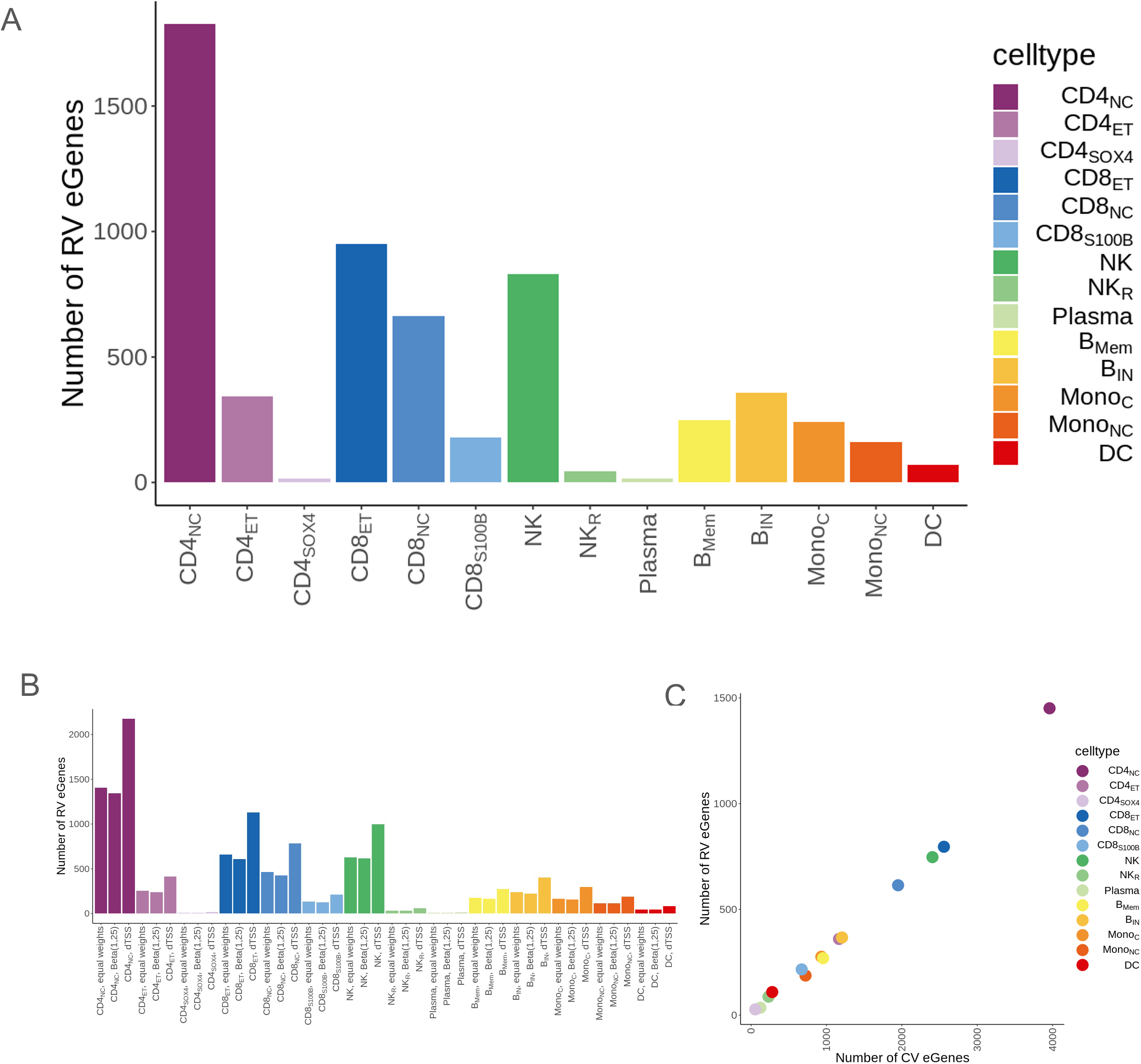
Rare variant eQTLs identified in OneK1K. A. Overview of RV signals (MAF<=5%) across 14 cell types (all *cis* variants collapsed). Bars represent the number of significant eGenes (FDR<5%). B. Effect of different weighting strategies in the set-based tests across all cell types. Compared methods are equal weights, Beta(1,25) distribution, weight based on the distance of each variant from the transcription start site (dTSS). C. Scatterplot of the number of number of significant eGenes with common variant signals (x axis) and rare variant signals (y axis) across the 14 cell types.

## DISCUSSION

Single-cell eQTL mapping has recently become an important tool for identifying the molecular underpinnings of genetic variation at a cellular level. However, current methods still present several limitations. The most commonly used methods, such as Matrix eQTL^40^, TensorQTL^38^, and QTLtools^56^ implement linear regressions, which cannot account for any structure between the samples. Therefore, they are limited to using pseudobulk approaches in unrelated samples, where single-cell counts are aggregated to a single value for each individual, gene, cell type^57^. This is typically done considering mean expression levels, which fail to model cell-to-cell variability and to exploit shared signals between cells from the same individual. Newer methods that directly model single-cell expression profiles exist but are limited in speed and scale and are currently only used as downstream interpretation tools rather than for discovering new genetic signals^15,16^.

Here, we introduced SAIGE-QTL, an efficient open-source tool implementing a Poisson mixed model. This tool overcomes these limitations by modelling single-cell profiles directly as discrete counts, accounting for both individual effects and relatedness between individuals, and is scalable and efficient, which allows fast genome-wide scans. Additionally, compared to existing methods, SAIGE-QTL implements not only single-variant tests but also set-based tests, which are better powered than single-variant tests for low-frequency variants.

We demonstrated that SAIGE-QTL is well calibrated for both the single-variant test for common variants and the set-based tests for rare and less frequent variants. As evaluated using the OneK1K dataset^7^, we have shown that eQTL mapping at the cellular level using SAIGE-QTL identified 48.4% more eGenes than the previous analyses using the pseudobulk approach. For about a third of the number of eGenes identified through common variant tests, we identified rare (MAF≤5%) variant eGenes through set-based tests, of which 21% were independent of common eQTL signals from the same genes. In addition, we identified 413 *trans* eQTLs for 390 genes in three representative cell types (CD4_NC_, B_IN_, Plasma).

SAIGE-QTL has existing limitations. First, the Poisson model is used to model the discrete read counts. While it has been recommended in the literature to use for scRNA-seq read counts, it is not necessarily the best model for all genes. Negative Binomial regression can be more appropriate to model read counts of genes with over-dispersion in scRNA-seq data. Second, as expected, the Poisson mixed model used for modelling single-cell read counts by SAIGE-QTL does not always have higher power for all genes than the pseudobulk approach in TensorQTL. Further investigation needs to be conducted to evaluate why some genes have much more significant p-values in pseudobulk analyses and vice versa. For example, the sparsity levels before and after pseudobulking may play a role as shown in **Supplementary Figure 8**. Third, SAIGE-QTL does not test for interactions of genotypes and other cell contexts, such as cell states, which has been previously reported to be critical for identifying dynamic eQTLs^15^. Finally, several approaches used by SAIGE-QTL to enhance computational efficiency are not directly applicable to random slope mixed models. These models can accommodate cells with diverse correlation structures instead of treating cells as repeated measurements. For example, read counts from cells may exhibit spatial correlation when leveraging spatial transcriptomics data. These limitations will be addressed next using the SAIGE-QTL framework.

## METHODS

SAIGE-QTL builds on the previously proposed SAIGE^22^ and SAIGE-GENE^23,24^ tools, with some key additions: i) the phenotype is modelled to follow a Poisson distribution, which is better suited for the single-cell expression count data; ii) random effect accounting for multiple cells per individual (in addition to the random effect accounting for relatedness between individuals); iii) improved computational efficiency for fitting Poisson mixed models with a random intercept; iv) ACAT-V^28^ test in addition to the Burden and SKAT for set-based tests for less frequent/rare genetic variants with multiple user-specified weights; v) the Cauchy method (ACAT-O^28^) for gene-level p-value combination to identify *cis*-eGenes; vi) *cis* window file as an input file to the open-access software to facilitate *cis* testing, and multi-gene implementation for genome-wide (*trans*) tests. The key steps are summarised below and details of our method can be found in the **Supplementary Note**.

### SAIGE-QTL

In a scRNA-seq study that sequenced *N* cells from *n* individuals, a Poisson generalised linear mixed model is used to model the read counts from the *jth* cell of the *ith* individual *y*_*ij*_ ~ *Poisson*(*μ*_*ij*_). Let the *X*_*cij*_ denote *p*_*c*_ cell-level covariates, such as cell states and cell-level expression PEER factors, including the intercept for the *jth* cell of the *ith* individual, and *X*_*di*_ denote *p*_*d*_ individual (donor)-level covariates, such as age, gender, genotype PCs, for the *ith* individual. Let *G*_*i*_ = 0,1,2 represent minor allele counts for the genetic variant of interest. The Poisson mixed model can be written as *μ*_*ij*_ = *exp*(*η*_*ij*_) and *η*_*ij*_ =*X*_*di*_ *α*_*d*_ + *X*_*cij*_*α*_*c*_ + *G*_*i*_*β* + *b*_*i*_, where *β, α*_*d*_, and *α*_*c*_ are the regression coefficients of the genotype *G*_*i*_, and the covariates *X*_*di*_ and *X*_*cij*_, respectively. Formulating the model using matrices and vectors, let ***Z*** denote the *N* × *n* design matrix with 1s and 0s to indicate which individual each cell belongs to. The model can also be written as ***η*** = ***X***_***c***_ ***α***_***c***_ + ***ZX***_***d***_ ***α***_***d***_ + ***ZGβ*** + ***Zb***_***d***_, where *η* is the *N* × 1 vector of *η*_*ij*_, ***X***_***c***_ is the *N* × (*p*_*c*_ + 1) matrix containing the cell-level covariates including the intercept, ***X***_***d***_ is the *n* × *p*_*d*_ matrix containing individual (donor)-level covariates, and the random effect ***b*** = ***Zb***_***d***_, where ***b***_***d***_ is the *n* × 1 vector of *b*_*i*_ and is assumed to follow multivariate Normal distribution 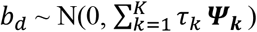 with ***Ψ***_***k***_ being the known *K* variance-covariance matrices, which can include the identity matrix to account for intra-individual variance of read counts across multiple cells and the genetic relationship matrix to account for sample relatedness, if any. We test the null hypothesis of no genetic association ***H***_**0**_ **: *β*** = **0** vs ***H***_**1**_ **: *β*** ≠ **0**.

#### Estimating the variance component and other model parameters (Step 1)

Similar to SAIGE^22^ and GMMAT^31^, in Step 1, SAIGE-QTL fit the null model ***η*** = ***X***_***c***_ ***α***_***c***_ + ***ZX***_***d***_ ***α***_***d***_ + ***Zb***_***d***_. The model parameters are estimated iteratively using the PQL^29^ method and the AI-REML algorithm^30^. At iteration ***q***, let 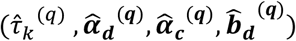, be estimated as 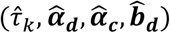, let 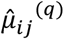 be the estimated mean of *y*_*ij*_, 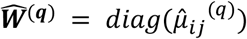, and let 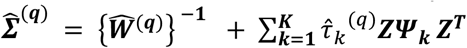 be an *N* × *N* matrix of the variance of working vector 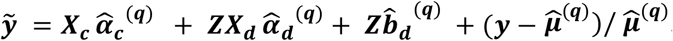. The PCG^32^ approach is used to calculate the product of 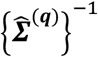 and an *N* × 1 vector at each iteration of model fitting, which is much more cost efficient than obtaining 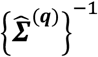 through matrix decomposition. Note that using the PCG approach, the product of 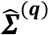 and an *N* × 1 vector needs to be calculated, which can be done with consecutive multiplications of the matrices ***Z***^***T***^, ***Ψ***_***k***_, **Z** and the *N* × 1 vector and does not require pre-computing and storing the *N* × *N* matrix ***ZΨ***_***k***_ ***Z***^***T***^. This reduces the computation time to calculate the product from *O*(*N*^2^) to *O*(*nN*) and is particularly efficient as *n* ≪ *N e*.*g*., when millions of cells are generated from hundreds or thousands donors in the study.

#### Estimating the variance ratios

A score test statistics for *H*_0_ : *β* = 0 is 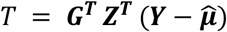, where *G* is the *n* × 1 genotype vector, *Y* is the *N* × 1 phenotype vector with read counts from cells, 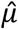 is the *N* × 1 vector with estimated mean of *Y* under *H*_0_. The variance of *T* is 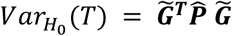, where 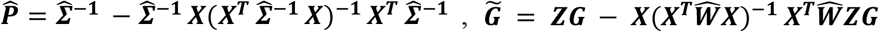, is the covariate-adjusted genotype vector, *X* is the *N* × (*p*_*c*_ + 1 + *p*_*d*_) matrix with all covariates including the intercept, and ***X*** = [***X***_***c***_ ***ZX***_***d***_]. For each genetic variant, given 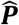, the calculation of 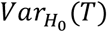 requires *O*(*N*^2^) computation. Similar to SAIGE^22^, BOLT-LMM^33^ and GRAMMAR-Gamma^34^, let 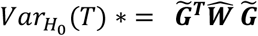, which does not incorporate random effects, and it has been shown previously that 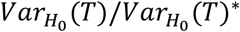 is approximately constant for all genetic variants with MAC ≥ 20 ^22,33,34,58^.

Here we estimate two variance ratios. First, similar to what has been used in the GWAS method fastGWA-GLMM^59^, we use the unadjusted (but mean-centred) genotype data, 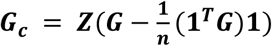, where **1** is a column vector of ones of size *n* × 1, to calculate 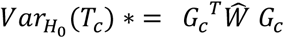 and then estimate the variance ratio 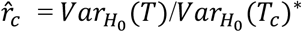. Second, to approximate the 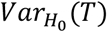 more accurately, we incorporate the random effects via 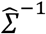 into the denominator to estimate the variance ratio 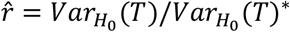, where 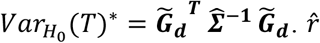 has lower variation than 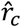 as previously demonstrated in SAIGE-GENE^23^. To further improve the computational efficiency, we use the donor-level covariates and intercept-adjusted genotype vector 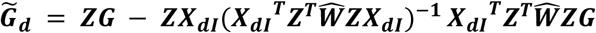 to compute 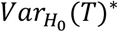 and then estimate 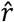. With pre-computed 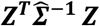 and 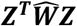, the computation cost of *Var*(*T*)* is *O*((1 + *p*_*d*_)*n*^2^) and *Var*(*T*_*c*_)* is *O*(*n*), respectively, instead of *O*(*N*).

We have shown that 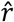 and 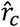 are approximately constant for all genetic variants (**Supplementary Note**). To estimate 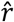 and 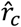, we calculate the variance ratios starting with 30 randomly selected genetic variants at an increment of 10 genetic variants until the coefficient of variation (CV) is smaller than 0.001 and calculate the mean of the variance ratios as 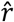 and 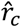, respectively.

#### Score test for single variants and set-based tests for rare variants (Step 2)

Using the estimated variance ratio 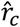, the variance-adjusted test statistic can be calculated as 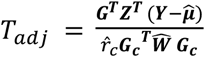, under the null hypothesis has mean zero and variance unity. For variants with p-value < 0.05, we use 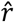 to re-calculate 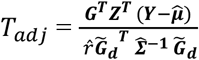. This strategy allows SAIGE-QTL to reduce the computation cost to calculate *T*_*adj*_ from *O*((1 + *p*_*d*_)*n*^2^) to *O*(*n*) for ~95% of genetic markers. In addition, it has been previously observed that the asymptotic normality assumption of the score test statistic *T*_*adj*_ leads to severe Type I error inflation for low-frequency and rare variants in the context of logistic mixed models with case-control ratios are unbalanced ^22–24^ and the modified Poisson mixed model when the event rate is low^58^. SPA^25^ has been used to successfully approximate *T*_*adj*_ for better controlled type I error rates. In SAIGE-QTL, we also implement and apply SPA to obtain better approximation of the distribution of *T*_*adj*_ to obtain more accurate p-values (**Supplementary Note**).

For eQTL analysis of rare variations due to the lack of power of single-variant tests, SAIGE-QTL conducts the set-based tests, including the Burden, SKAT^27^, and ACAT-V^28^ tests, Test statistics of the Burden and SKAT^63^ tests can be constructed based on score statistics from the marginal model for individual variants in the testing set and test statistics of the ACAT-V^28^ test can be constructed using p-values from the marginal model for individual variants. Following what has been proposed in SAIGE-GENE+^24^, to reduce the data sparsity due to ultra-rare variants, before testing each variant set, ultra-rare variants with MAC ≤ 20 are collapsed in SAIGE-QTL to a pseudo-variant and then the pseudo-variant is tested together with all other variants with MAC > 20. In addition, multiple functional annotations and maximum minor allele frequency cutoffs are allowed to be incorporated into the set-based tests to improve power and p-values are combined using the minimum P value or Cauchy combination method^28^.

#### Calculating gene-level p-values for identifying cis-eGenes (Step 3)

In Step 3, the gene-level p-value for each gene is calculated by combining the variant-level p-values of common variants (MAF>5% is used in the OneK1K study) using the ACAT-V^28^ test based on the Cauchy combination as recently suggested in the linear mixed model method for eQTL mapping, APEX^36^. We assume that there are *M* genetic variants in the *cis*-region, let 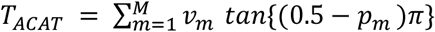, where *p*_*m*_ is the p-value for the *mth* variant, *v*_*m*_ is the weight for the *mth* variant and let 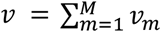. The p-value of *ACAT* is approximated by *p*_*ACAT*_ ≈ 1/2 − {*arctan* (*T*_*ACAT*_ /*v*)}/π. It is a computationally efficient approach to combine p-values, which is robust to the correlation between individual p-values^28^. It provides highly concordant gene-level p-values compared to those obtained using the Beta approximation method^37,38^ (**Supplementary Figure 2**), but is much faster as it does not require any permutations to be performed. It also allows for incorporating variant-level weights, *e*.*g*., distance to transcription start site, to improve the power of detect *cis*-eGenes.

#### OneK1K analysis

We consider the OneK1K dataset as it is available from the original publication^7^. This includes genotypes (from SNP array data) and scRNA-seq from 14 different immune cell types for 1,267,758 peripheral blood mononuclear cells (PBMCs) and 982 individuals of European ancestry from the Tasmanian Ophthalmic Biobank in Tasmania, Australia. For eQTL mapping with both SAIGE-QTL and TensorQTL, we considered the same covariates used for eQTL mapping in the original paper: sex and age of the individuals, six genotype PCs and 2 PEER factors calculated from gene expression from each cell type separately. Since the original study used pseudo-bulk aggregate counts, the PEER factors are defined per individual, not cell, so we kept those values here (*i*.*e*., all cells for a given individual were assigned the same PEER factor value). As used in the original publication^7^, the SCTransform v1 function in the Seurat R package^60^ was used to normalise read counts by total reads and account for batch and mitochondrial gene percentage, while still returning Poisson counts. The corrected read counts were used as input for Step 1 in SAIGE-QTL to fit null Poisson mixed models for each gene in different cell types.

For each gene in each cell type, we calculated the gene-level p-value in Step 3 based on all genetic variants within a 1Mb window around the gene body with MAF>5%. No weights are incorporated in the Cauchy combinations for a fair comparison with the results using TensorQTL.

Cell type name abbreviations: CD4_NC_: CD4^+^ naïve and central memory T cells, CD4_ET_: CD4^+^ T cells with an effector memory or central memory phenotype, CD4_SOX4_: CD4^+^ T cells expressing SOX4, CD8_ET_: CD8^+^ T cells with an effector memory phenotype, CD8_NC_: CD8^+^ naïve and central memory T cells, CD8_S100B_: CD8^+^ T cells with expression of S100B, NK: natural killer cells, NK_R_: natural killer recruiting cells, Plasma: Plasma cells, B_Mem_: memory B cells, B_IN_: immature and naïve B cells, Mono_C_: classical monocytes, Mono_NC_: nonclassical monocytes, DC: dendritic cells.

#### Data simulation for type I error evaluation

We first selected three representative cell types (CD4_NC_, B_IN_ and Plasma) with high, medium, and low abundance (**Supplementary Table 1**). Next, we randomly selected 50 genes with low (< 20%), medium (45 ~ 55%) and high (> 80%) sparsity levels, defined as the proportion of zero read counts, in each cell type separately (**Supplementary Table 3**). For each gene, we fit a null Poisson mixed model on the single-cell counts for the selected genes using the same covariates as described in the “OneK1K analysis” subsection above. Once the model is fitted, before running Step 2 for association tests, we permuted individual IDs in the genotype file to disrupt the possible association between genotype and gene expression. With each permutation, single-variant association tests on 405,663 genetic variants on chromosome 1 with MAF>5% were performed (Step 2a). Permutation was repeated for 250 times to obtain a sufficient number of tests for estimating empirical type I error rates at different *α* levels (**Supplementary Table 4**). ACAT-V tests were then performed for each gene region on chromosome 1 to estimate the empirical type I error rates at different *α* levels for Step 3 as shown in **Supplementary Table 5**.

The set-based tests (Step 2b) were performed for rare/less frequent genetic variants with MAF ≤5% in each gene region on chromosome 1 for three randomly selected genes from each sparsity group in three cell types. Two sets of weights were used, 1) equal weights, where each variant is assigned the same weight, 2) weights following a Beta(1,25) distribution, as commonly used in rare variant association tests. The tests were repeated for 10 times with permuted individual IDs and the Q-Q plots shown in **Supplementary Figure 5**.

#### CD4_NC_ cell subsetting

For the subsetting analysis (**Figure 1D**), we considered the most abundant cell type from OneK1K, CD4_NC_ (CD4 naive and central memory T cells). For each percentage level of subsetting (1, 5, 10, 20 and 50%), we subsetted the number of total cells to that percentage, while retaining the cell to donor ratio for each individual included. Next, we re-mapped *cis* eQTLs using SAIGE-QTL for each set, maintaining all other settings (covariates included, *cis* window size). Only genes expressed in at least 1% of the cells were tested, so those numbers decreased for smaller sets. Pseudobulking for TensorQTL was performed post subsetting, by considering the mean expression for each gene, cell type and individual. These values were then logged prior to eQTL mapping, to mirror the approach used in the OneK1K paper^7^.

#### Mendelian Randomization analysis

To identify genes whose expression levels are associated with human complex disease risk, we utilised the Summary-data–based Mendelian randomization (SMR)^61^ framework and used the top associated *cis*-eQTL of the gene as an instrumental variable (IV). We input the eQTL summary statistics of OneK1K cohort from SAIGE-QTL and four GWAS summary statistics of autoimmune diseases: Crohn’s disease (CD)^43^, inflammatory bowel disease (IBD)^43^, rheumatoid arthritis (RA)^44^, and systemic lupus erythematosus (SLE)^45^. We also ran the SMR tool using the eQTL summary obtained from Matrix eQTL^40^ for the same 14 cell types as a comparison. A HEIDI (HEterogeneity In Dependent Instruments) test implemented in the SMR framework was further performed to distinguish the pleiotropy/causality from linkage. The genes that are not rejected by the HEIDI test indicate that the gene expression levels are associated with the disease risk due to pleiotropy/causality at one shared genetic variant.

For each cell type, we calculated the q-values of each SNP-gene pair (the SNPs within a 1Mb window around the gene body) and only tested the genes whose top associated *cis*-eQTL pass the threshold of FDR<5%, to match what we used throughout the study (the thresholds used for each method and cell type are provided as **Supplementary Table 8**). Specifically, for each cell type we considered the weakest eGene still under the threshold of FDR<5%. Then, we considered the smallest p-value for that gene, across the *cis* SNPs tested (note that this necessarily results in the number of genes tested being different between SAIGE-QTL and Matrix eQTL). The significance of the SMR test was determined at p-value from SMR < 0.05 / M, where M is the total number of genes tested in each cell type.

#### Trans eQTL mapping

For each of the three representative cell types (CD4_NC_, B_IN_ and Plasma), considering the large number of tests and limited power for detecting *trans*-eQTLs with low frequency, we conducted genome-wide single-variant tests for all common variants with MAF >10%. We define *trans* signals as those outside of a 2Mb window around the gene for same-chromosome, as well as all inter-chromosome associations. Additionally, we exclude the MHC region on chromosome 6.

We report results at two different significance levels, *p* < 5 × 10^−6^ (**Supplementary Figure 13**) and *p* < 5 × 10^−8^ (**Figure 3**). At *p* < 5 × 10^−8^, we identified 223 *trans* eQTLs for CD4_NC_, 124 for B_IN_, 690 for Plasma (211, 120 and 69 unique genes, respectively), for a total of 413 *trans* eQTLs for 390 unique genes across all 3 cell types. At *p* < 5 × 10^−6^, we identified 47,210 *trans* eQTLs for CD4_NC_, 33,936 for B_IN_, 13,677 for Plasma (12,693, 10,477 and 5,460 unique genes, respectively), for a total of 94,795 *trans* eQTLs for 13,159 unique genes across all 3 cell types.

### Rare variant testing

When testing for the effects of rare variation (MAF≤5%) on single-cell gene expression, we used the Burden, SKAT and ACAT-V tests, and used the Cauchy approach to combine p-values. We also considered three sets of weights, 1) equal weights, where each variant is assigned the same weight, 2) weights following a Beta(1,25) distribution, as commonly used in rare variant association tests, and 3) weights based on a variant’s distance from the gene’s transcription start site (TSS). For the latter, we followed the procedure outlined by the APEX authors^36^, whereby “each variant received a weight proportional to e^-γ|d|^ where d is the number of base pairs between the variant and TSS and γ = 1e-5. Additionally, in order to determine the independence of these signals from common variants’ effects we performed conditional analysis, conditioning on all common variant eQTL signals at FDR<5%, across all five rounds of conditional analysis (**Supplementary Figure 15**).

## Data Availability

The OneK1K single-cell RNA-seq and genotype data were available via Gene Expression Omnibus (GSE196830).
Summary statistics for the eQTLs in this study can be found on Zenodo

https://zenodo.org/records/10811106

## Code availability

SAIGE-QTL is implemented as an open-source R method available at: https://github.com/weizhou0/qtl

Code to reproduce all analyses and figures included here can be found at: https://github.com/annacuomo/SAIGE_QTL_analyses

## Data availability

The OneK1K single-cell RNA-seq and genotype data were available via Gene Expression Omnibus (GSE196830).

Summary statistics for the eQTLs in this study can be found on Zenodo: https://zenodo.org/records/10811106.

GWAS summary stats for the MR analysis were downloaded from: i) CD and IBD GWAS were downloaded from IBD Genetics Consortium website (www.ibdgenetics.org) from Liu et al. Nature Genetics. 2015; ii) RA GWAS summary was downloaded from GWAS Catalog (study ID: GCST90132223) in Ishigaki et al. *Nature Genetics*, 2022 (https://ftp.ebi.ac.uk/pub/databases/gwas/summary_statistics/GCST90132001-GCST90133000/GCST90132223/GCST90132223_buildGRCh37.tsv.gz); iii) SLE GWAS summary was downloaded from GWAS Catalog (study ID: GCST90018917) in Sakaue et al. *Nature Genetics*, 2021 (https://ftp.ebi.ac.uk/pub/databases/gwas/summary_statistics/GCST90018001-GCST90019000/GCST90018917/GCST90018917_buildGRCh37.tsv.gz).

## ACKNOWLEDGEMENTS

W.Z. was supported by the National Human Genome Research Institute of the National Institutes of Health under award number K99/R00HG012222. A.C. was supported by an EMBO Long Term Postdoctoral Fellowship (EMBO ALTF 424-2022). B.M.N. is supported by the Novo Nordisk Foundation (NNF21SA0072102) and R01MH101244. BMN and GC are supported by R37MH107649. B.M.N. is a member of the scientific advisory board at Deep Genomics and Neumora. M.D. is a founder of Maze Therapeutics.

## Supplementary Figures

**Suppl. Fig. 1:**
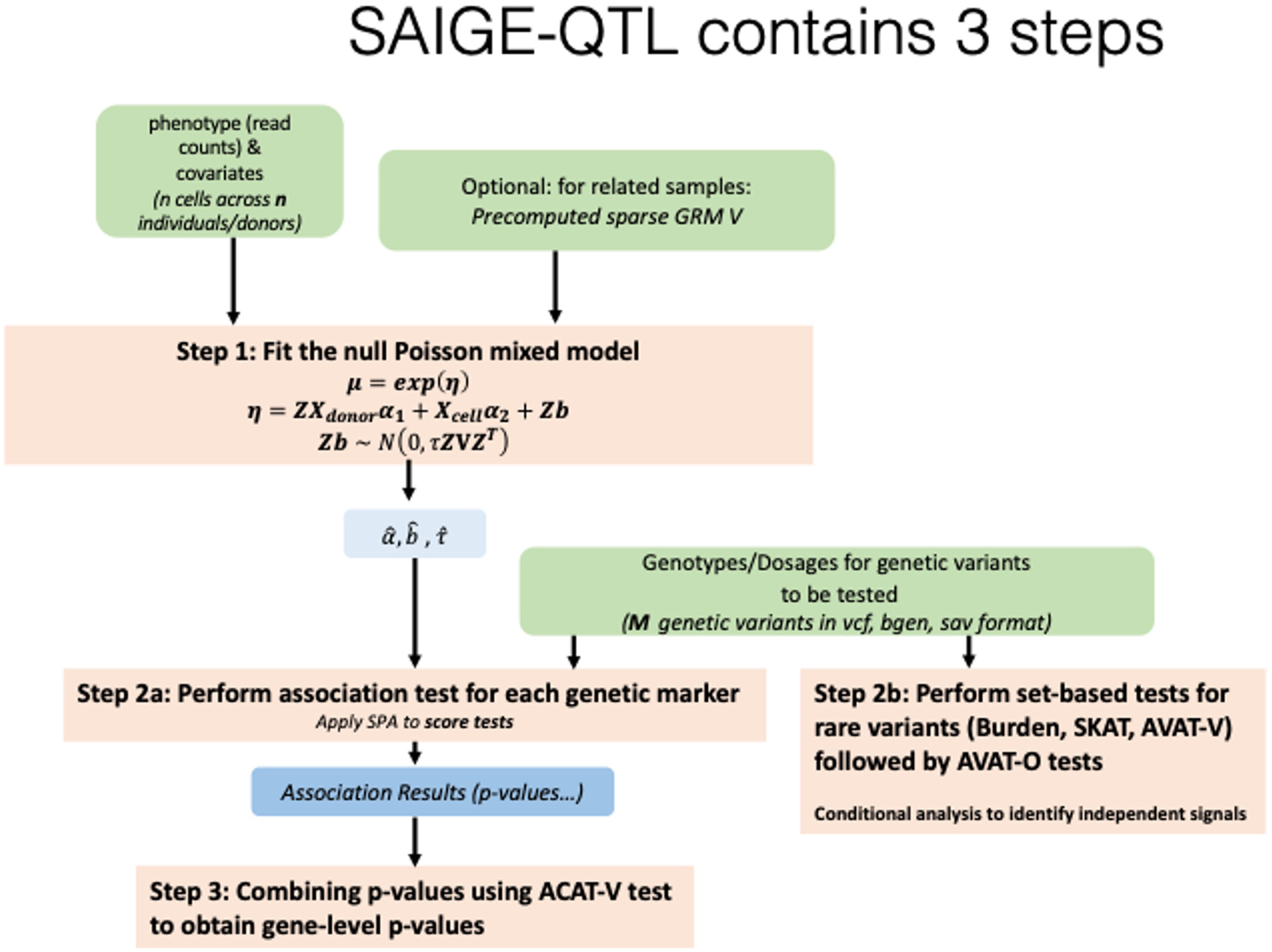
Method overview of SAIGE-QTL. Key steps, input arguments and parameters estimated are specified for our SAIGE-QTL method. Details can be found in the **Methods** and **Supplementary Note**.

**Suppl. Fig. 2:**
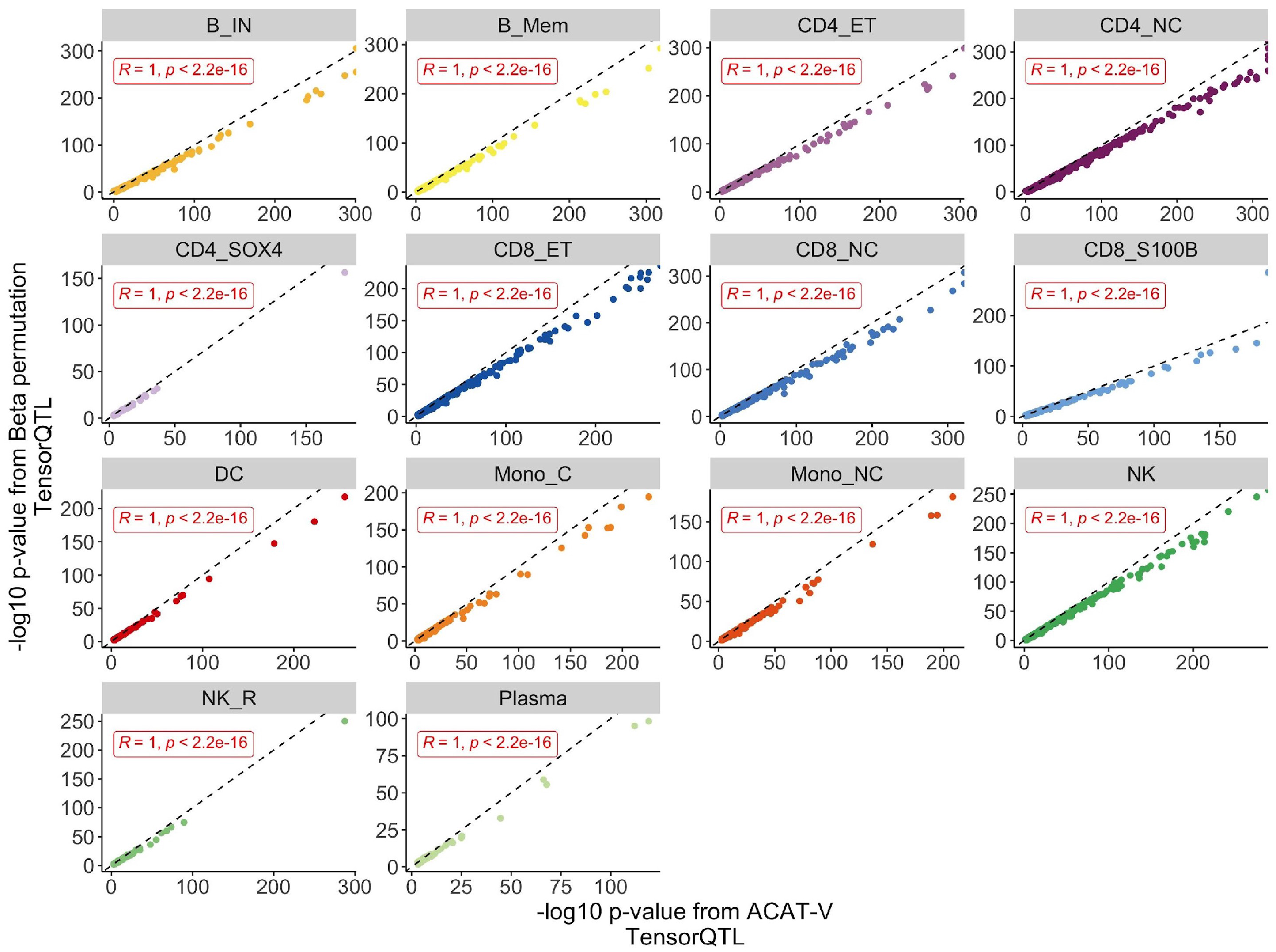
Concordance of gene-level p-values ac using the Beta permutation and the Cauchy combination methods. Scatterplots showing the concordance of negative log10 gene-level p-values calculated using different methods. Both axes show results using the nominal p-values from TensorQTL, but gene-level summary p-values are calculated using ACAT-V (Cauchy method, as we propose here, x axis) vs using the Beta approximation method used typically in TensorQTL (y axis).

**Suppl. Fig. 3:**
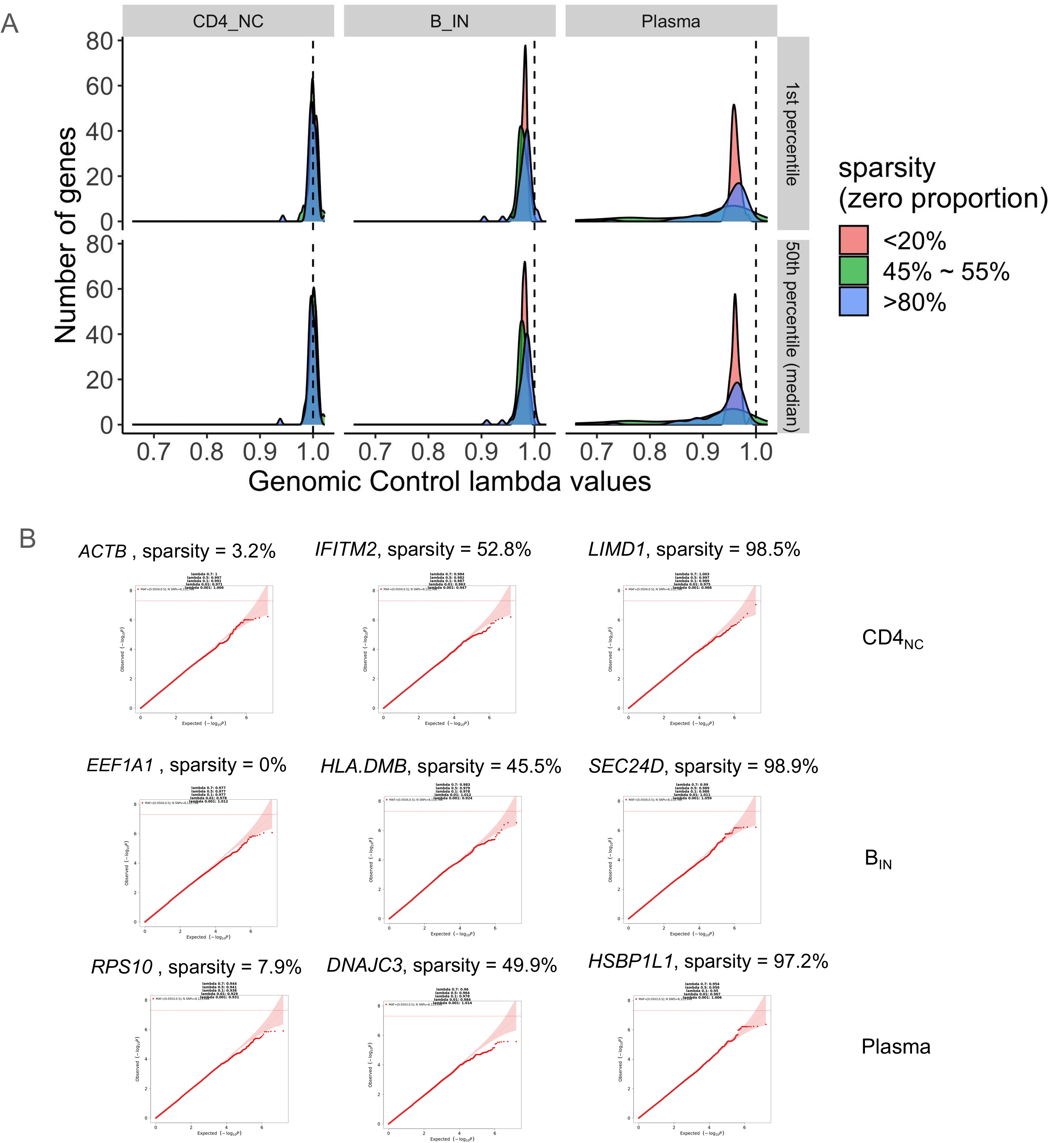
Evaluations of p-value calibration of SAIGE-QTL on common variants (MAF > 5%) from simulation studies under the null hypothesis. A. Genomic control lambda values of single-variant association tests. B. Quantile-quantile plots of 3 randomly selected genes from each sparsity group and cell type (results from chromosome 1 with 10 times of permutations are plotted). 50 genes were randomly selected from each sparsity group per cell type and single-variant association tests across chromosome 1 were conducted for the single-cell expression read counts of each gene.

**Suppl. Fig. 4:**
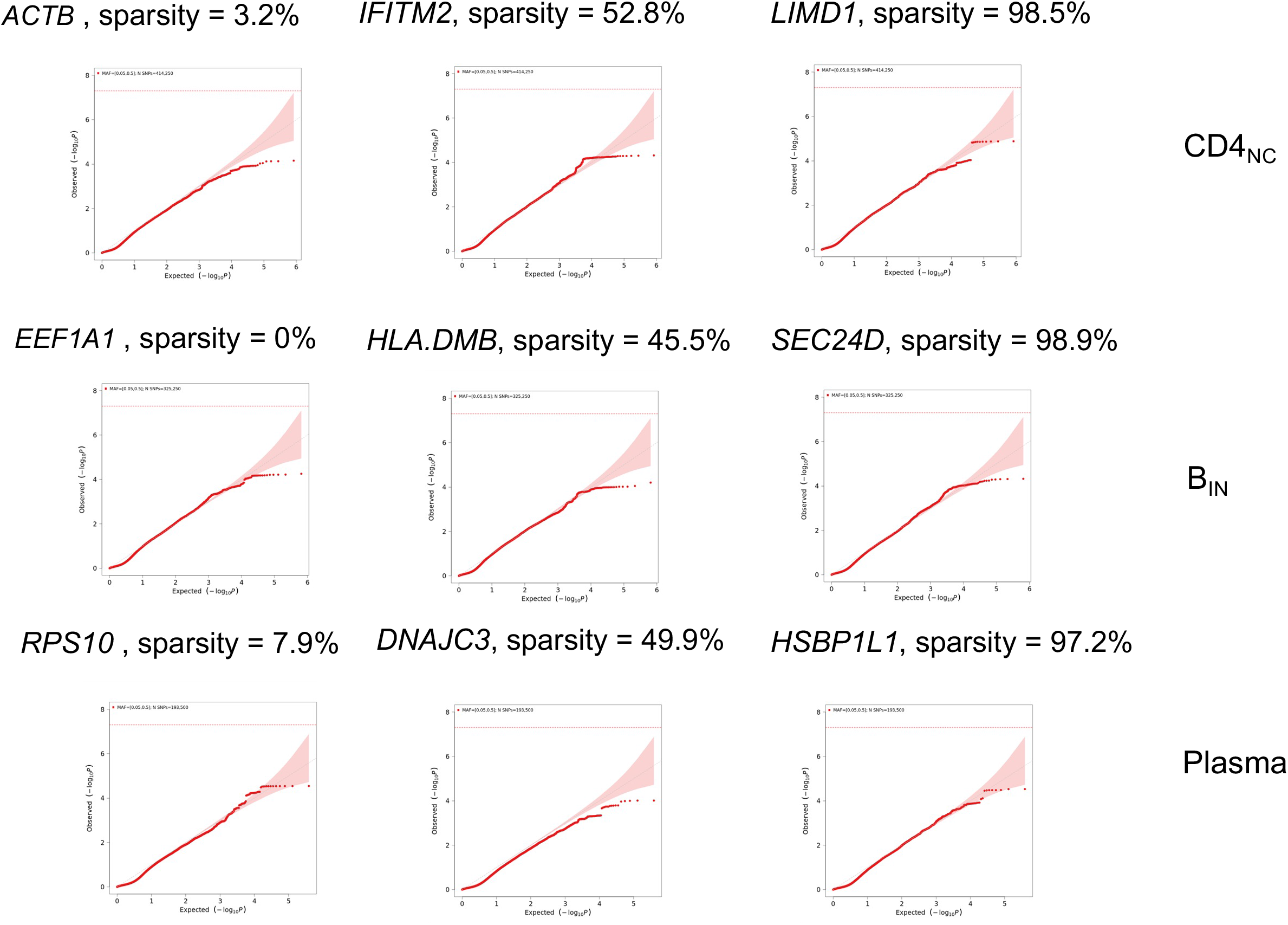
Quantile-quantile (QQ) plots of ACAT-V tests of common variants (MAF > 5%) on each gene region on chromosome 1 in simulation studies under the null hypothesis. Genes were randomly selected from each sparsity group (low, intermediate, high) per cell type and single-variant association tests across chromosome 1 with 250 times of permutations were conducted for the single-cell expression read counts of each gene (**Suppl. Fig. 3B**). For a *cis* window up and downstream of each gene body (+/−1Mb) on chromosome 1 the ACAT-V test is performed to obtain a gene-level p-value. Three representative cell types (Plasma: least abundant, naive and immature B cells, B_in_: intermediate, and CD4 positive naive and central memory T cells, CD4_NC_: most abundant) are shown.

**Suppl. Fig. 5:**
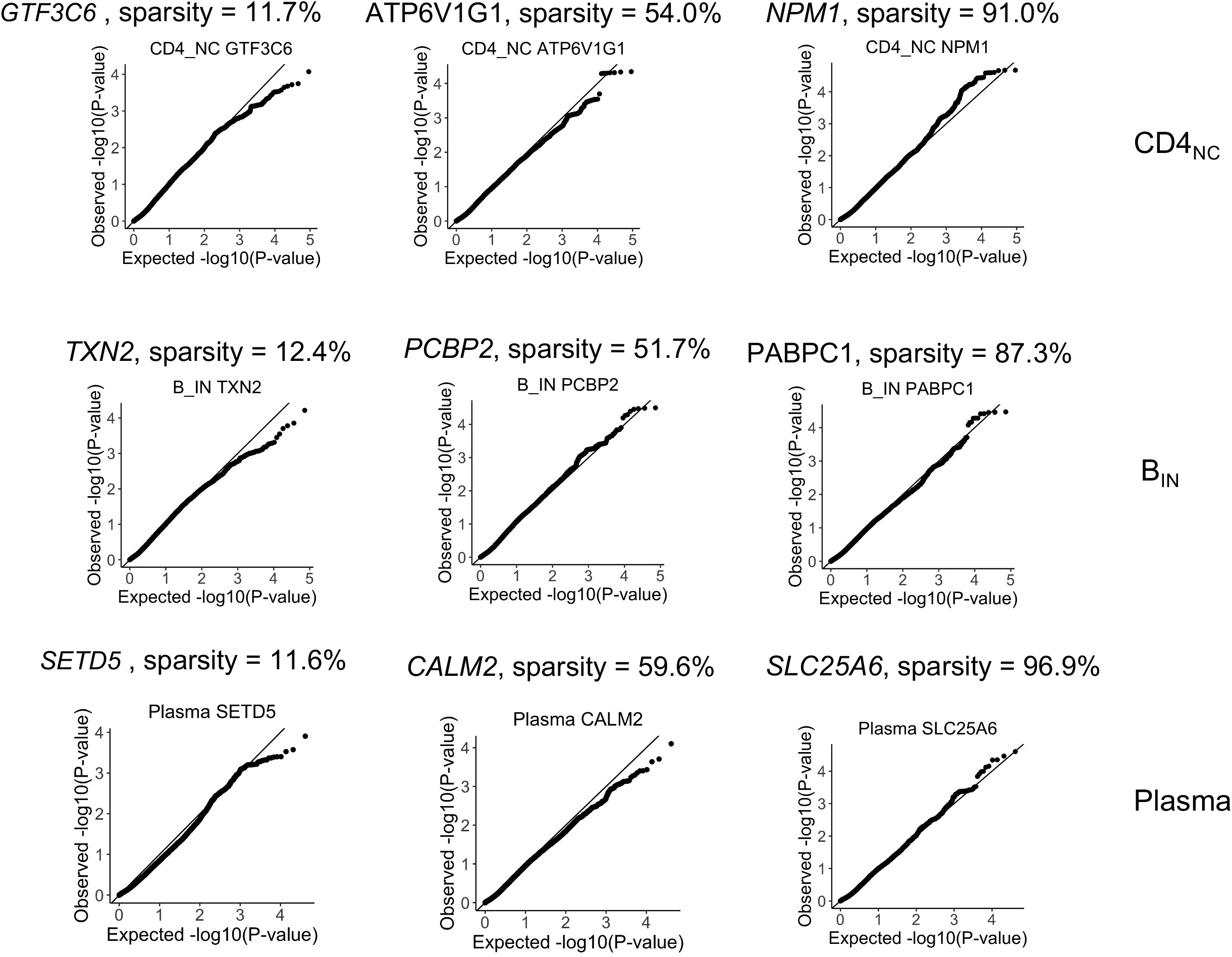
Quantile-quantile (QQ) plots of set-based tests of rare variants (MAF ≤ 5%) on each gene region on chromosome 1 in simulation studies under the null hypothesis. For three representative cell types (=rows, Plasma: least abundant, naive and immature B cells, B_in_: intermediate, and CD4 positive naive and central memory Tcells, CD4_NC_: most abundant), shown are the p-values for rare variants (MAF <= 5%) from simulation studies under the null hypothesis that there is no eQTL effects on gene expression across representative genes of varying sparsity (=columns, specific to each genes, low to high sparsity from left to right).

**Suppl. Fig. 6:**
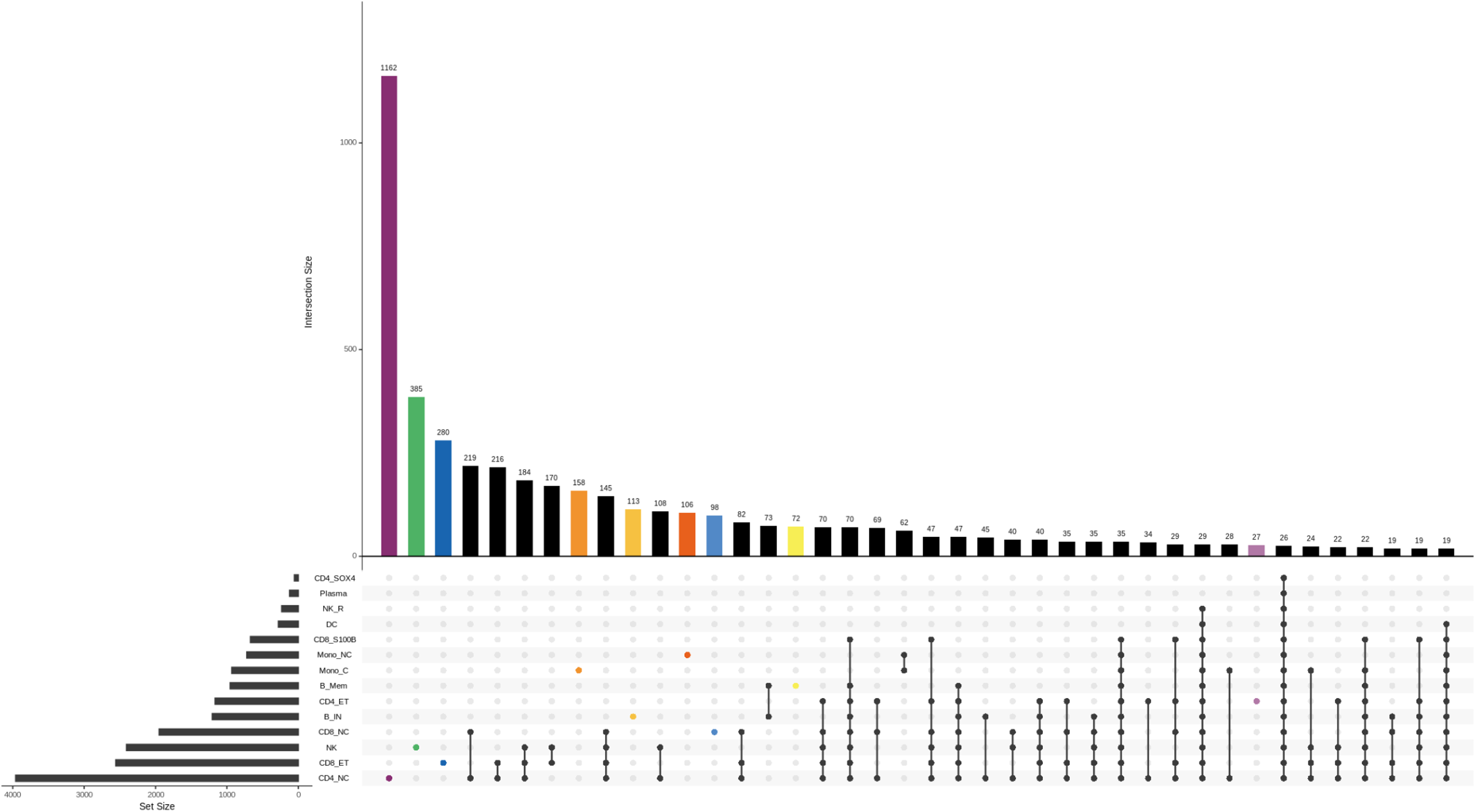
cell type-specific eQTLs. Upset plot showing the cell type specificity of significant eGenes (FDR<5%) identified by SAIGE-QTL in the OneK1K dataset. Coloured bars represent the number of eQTLs only found in a specific cell type, while black bars represent shared signals across two or more cell types. Values >10 are included. The UpSetR package in R was used for this figure.

**Suppl. Fig. 7:**
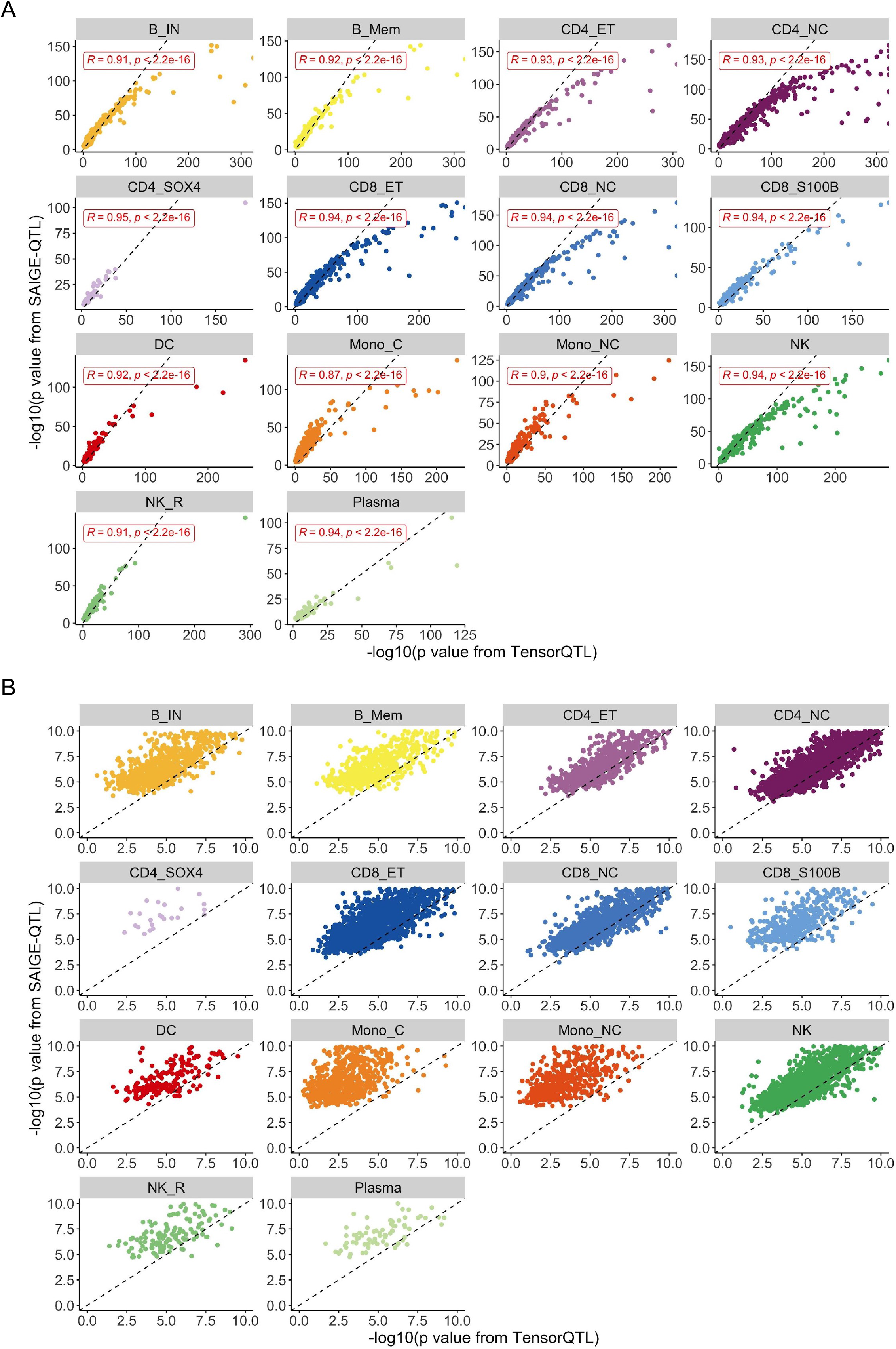
p-value concordance between SAIGE-QTL and TensorQTL. Scatter plots of negative log10 p-values from TensorQTL (x axis) and SAIGE-QTL (y axis) are shown for each of the 14 cell types. Top variants in eGenes identified in OneK1K (FDR < 5%) are included. A. All variants, with correlation coefficients specified. B. As in A, but zooming in to x and y axes values between 0 and 10.

**Suppl. Fig. 8:**
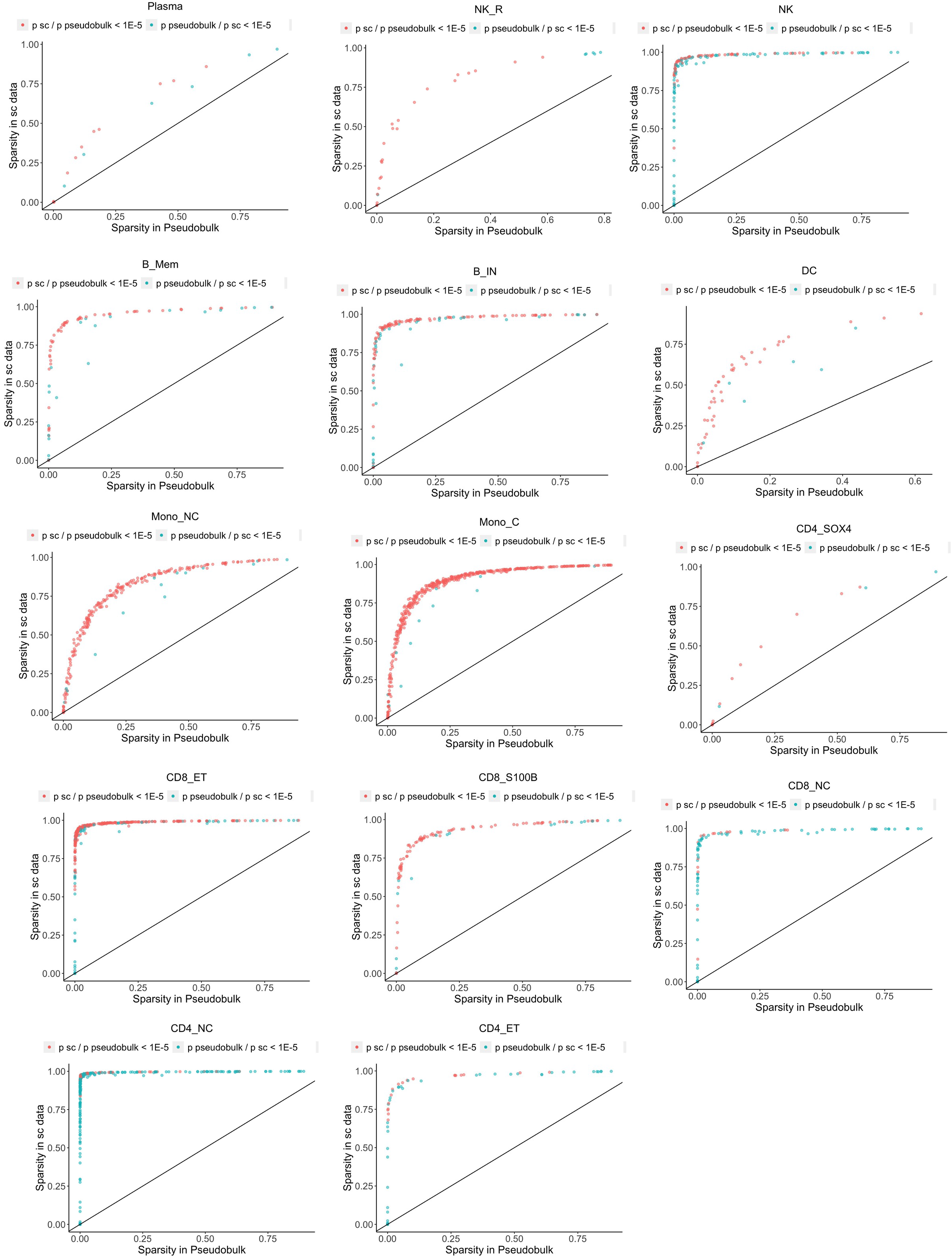
Relationship between sparsity levels and the nominal p-values of sc-eQTL using SAIGE-QTL and pseudobulk eQTL using TensorQTL. Sparsity level across cells (y axis) is plotted against sparsity level across donors (x axis) for top variants in eGenes with FDR < 0.05 that have different p-values in the two analyses. If the top variants are different identified by SAIGE-QTL and TensorQTL, the ones by TensorQTL are included. Each plot represents the results for each of the 14 cell types. Variants with more

**Suppl. Fig. 9:**
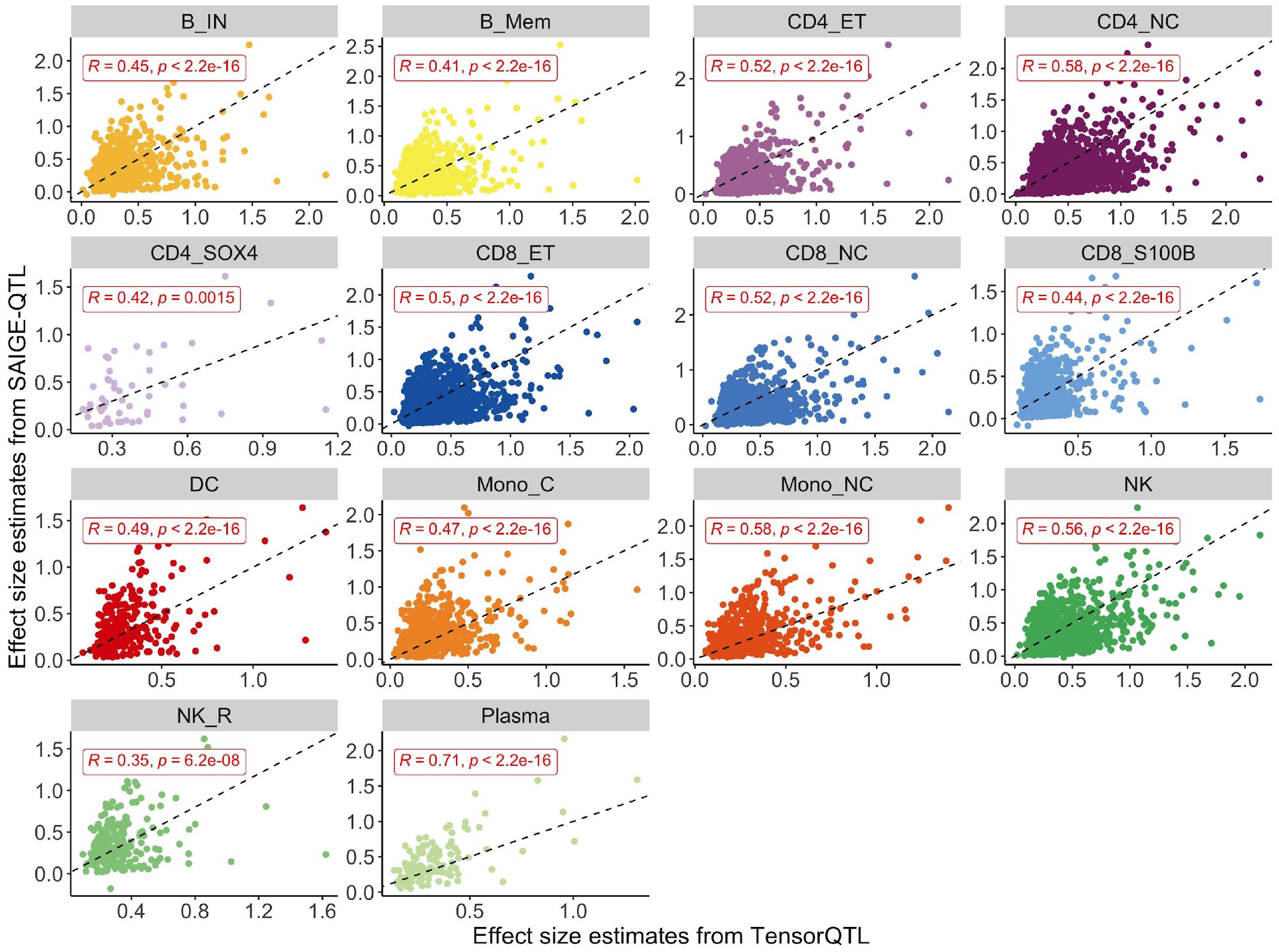
Effect size concordance between SAIGE-QTL and TensorQTL. Scatter plots of estimated effect sizes from TensorQTL (x axis) and SAIGE-QTL (y axis) are shown for each of the 14 cell types, and correlations are specified. Top variants in SAIGE-QTL results in each eGene identified by either of the two methods are included.

**Suppl. Fig. 10:**
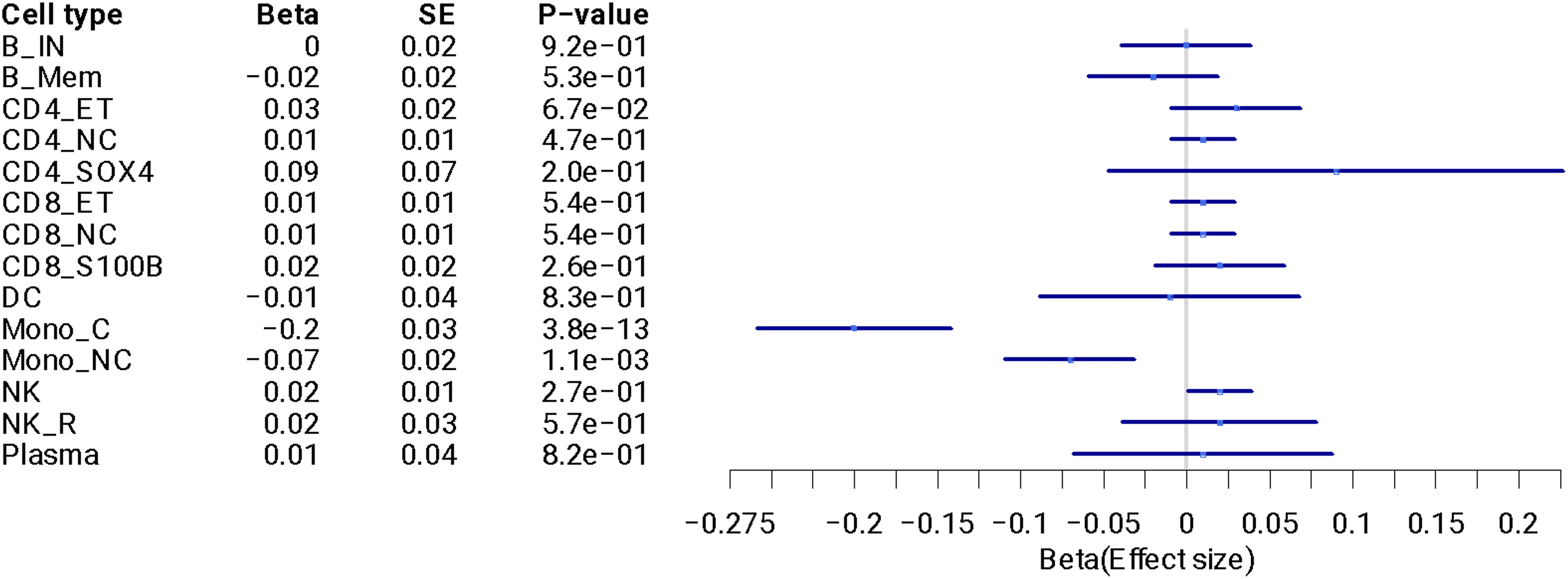
Forest plot of effect sizes of the top cis-eQTL variant rs1375493 in gene *ITGA4* across 14 cell types. For the significant eQTL identified for *ITGA4* and shown in Figure 2B, forest plot of effect sizes across all cell types, highlighting that this is a monocyte-specific signal.

**Suppl. Fig. 11:**
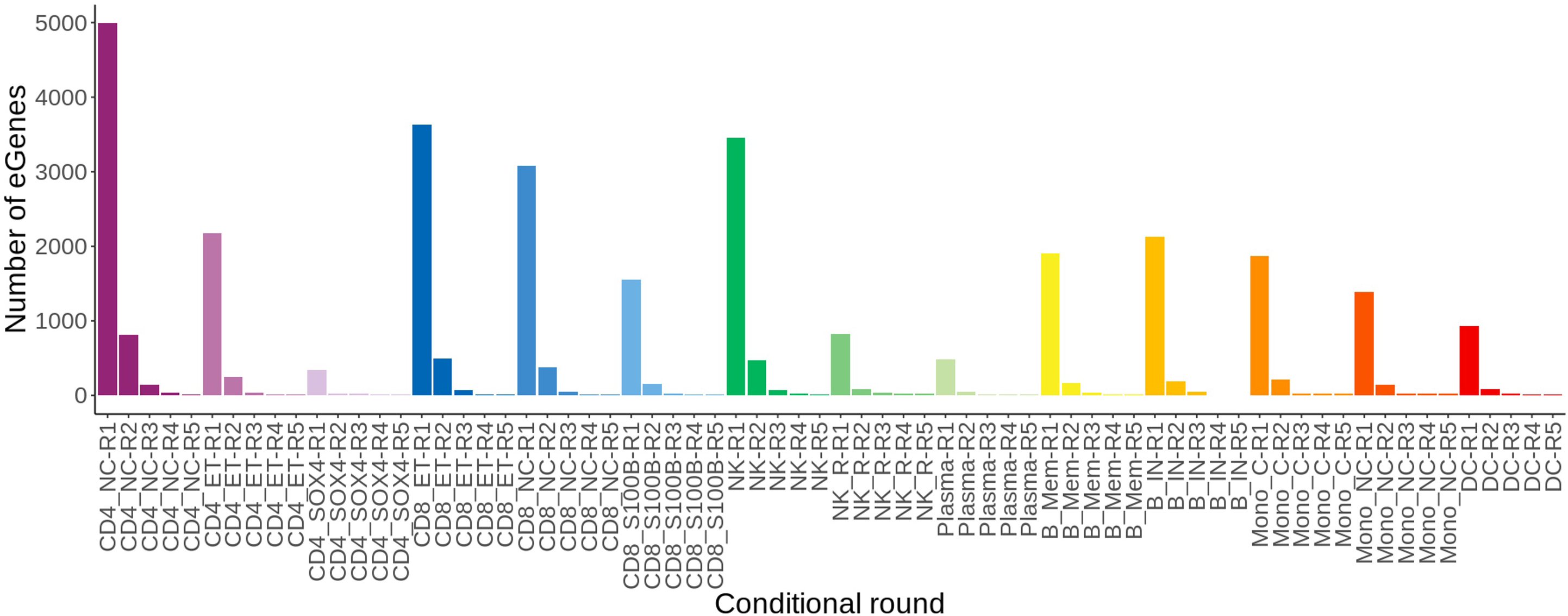
Common variants conditional analysis results. For each of 14 cell types (represented by the different colours), bars represent the number of significant eGenes (FDR<5%) identified for each of 5 rounds of conditional analysis (R1 to R5).

**Suppl. Fig. 12:**
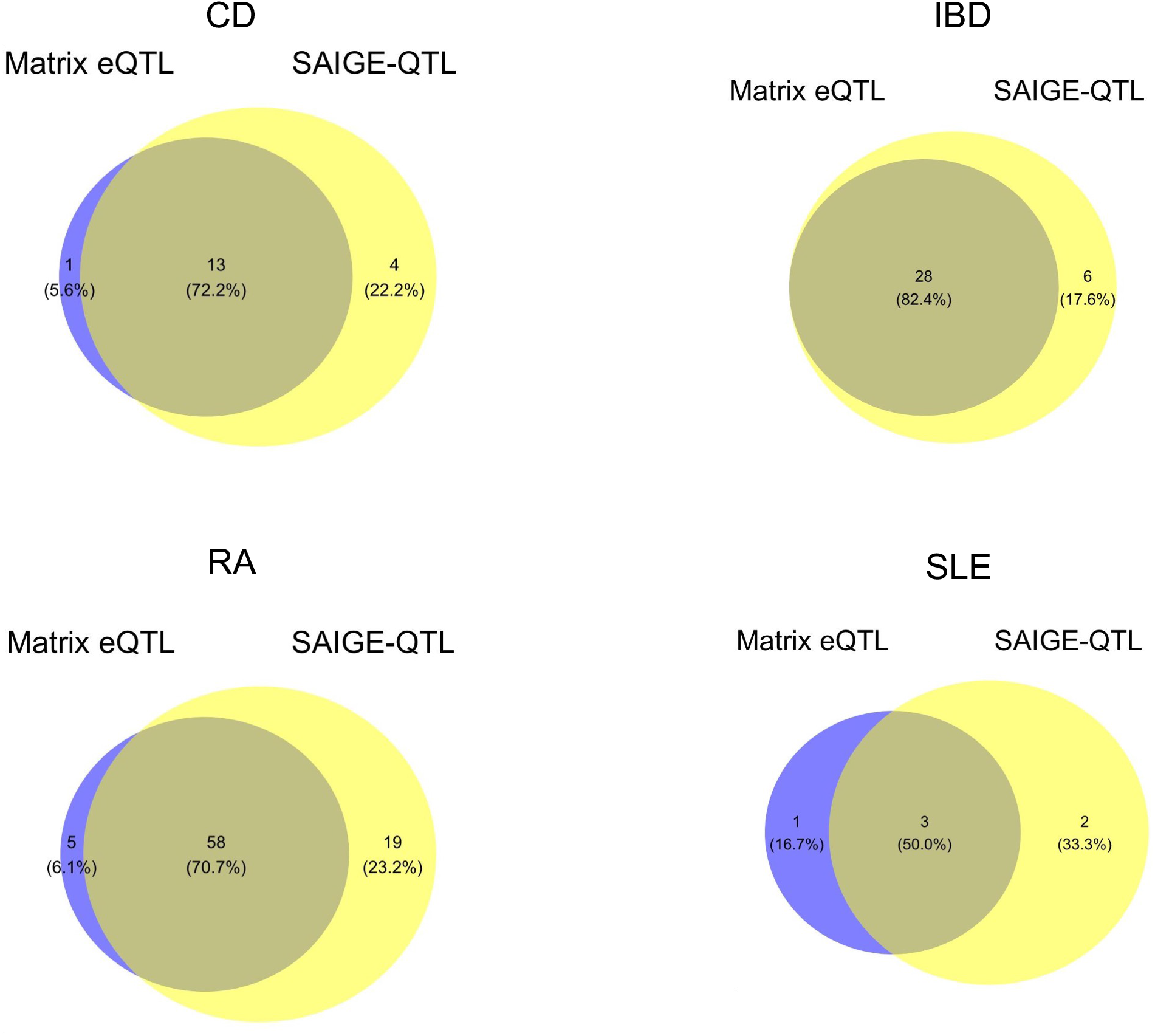
Number of loci identified in any cell type by SMR using eQTLs identified by the two methods. Venn diagrams representing the overlap of significant summary-based Mendelian Randomisation (SMR) results identified based on eQTLs detected using each of the two methods, for the three immune-mediated diseases considered. Similar plots for gene-cell type combinations are shown in the main figure (Fig 2C). CD: Crohn’s disease, IBD: inflammatory bowel disease, RA: rheumatoid arthritis, SLE: systemic lupus erythematosus.

**Suppl. Fig. 13:**
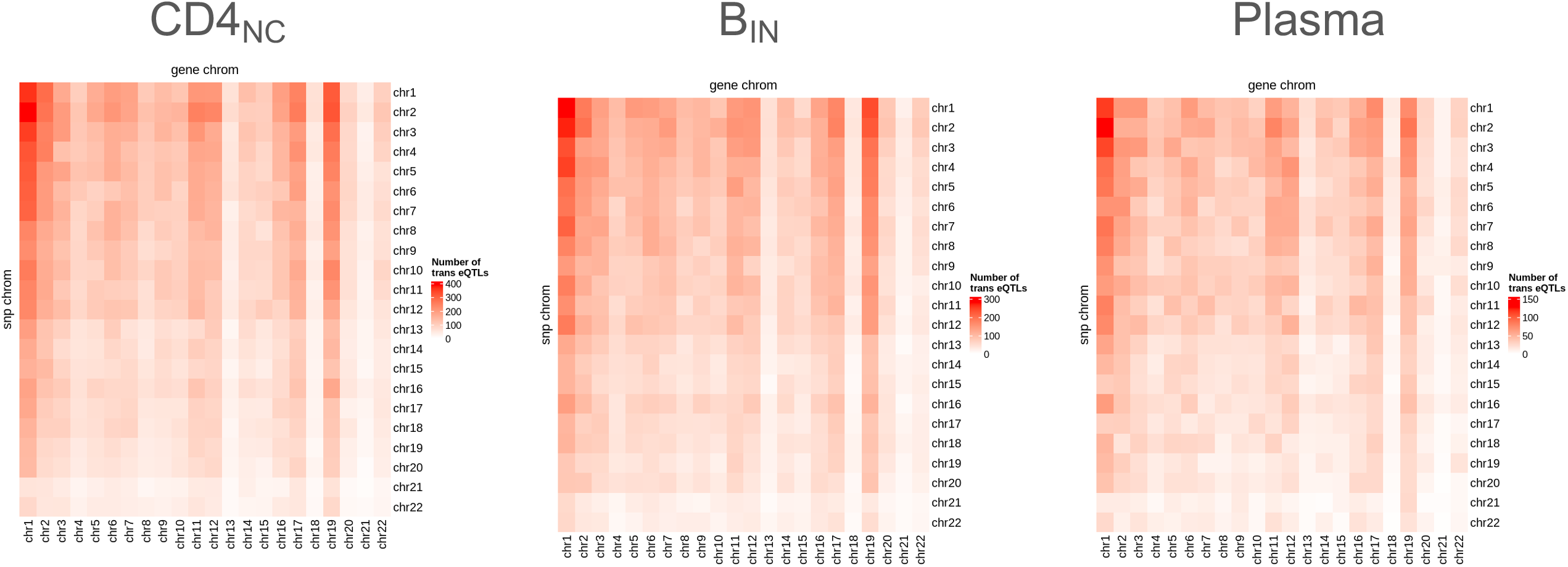
Trans eQTL effects across three representative cell types. Same as main figure (Fig. 3a), but considering all associations with p-value < 5e-6. For three representative cell types (Plasma: least abundant, naive and immature B cells, B_in_: intermediate, and CD4 positive naive and central memory Tcells, CD4_NC_: most abundant), heatmaps where the colour intensity represents the number of significant independent trans eQTLs. Rows represent the chromosome the variant is on, columns the gene chromosome.

**Suppl. Fig. 14:**
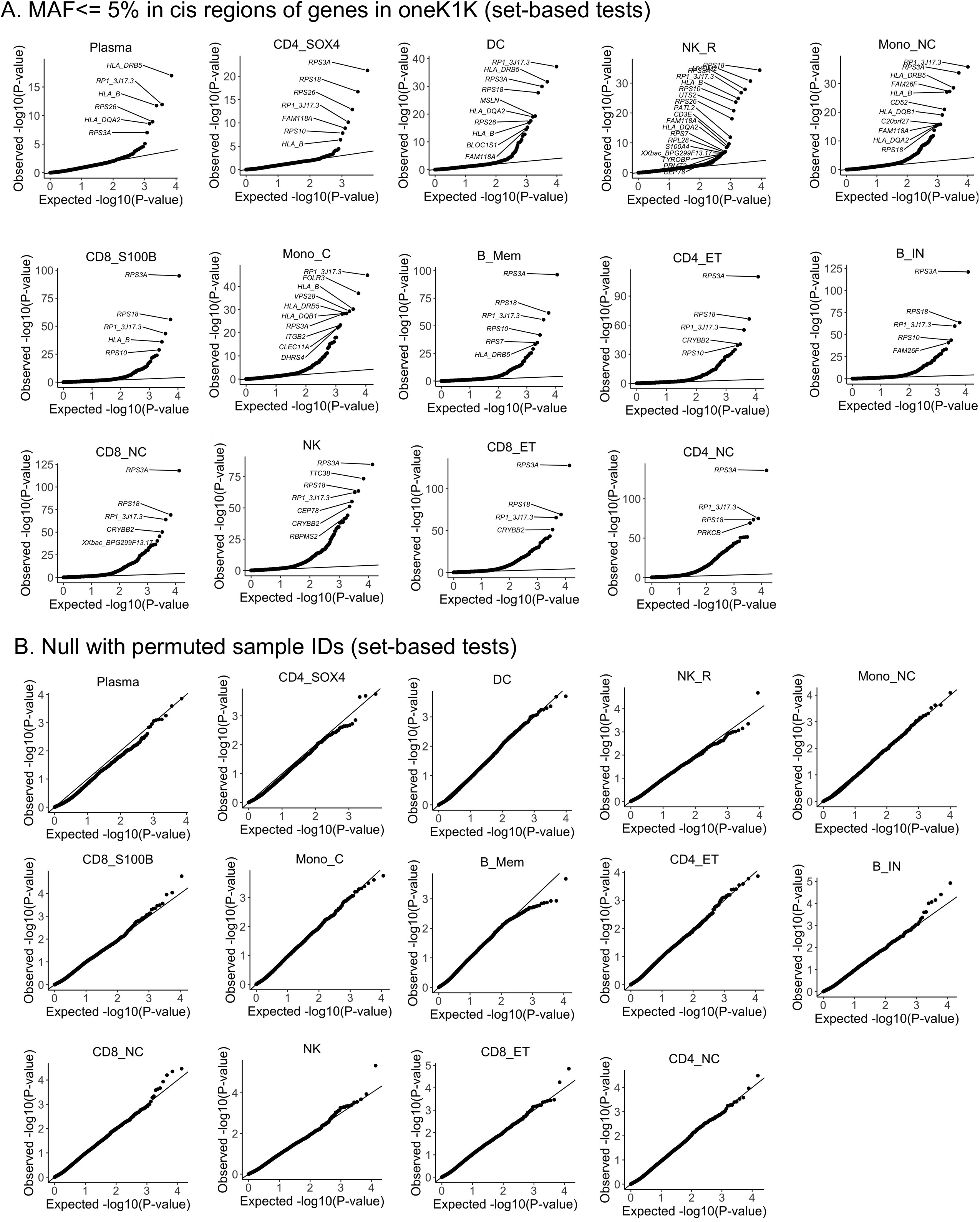
QQ plots of set-based test p-values for rare variants (MAF <= 5%) in OneK1K. A. All genes (expressed in >= 10% of donors) across 14 cell types. B. As in A, but after permutating genotypes to remove association signal to assess calibration of our test.

**Suppl. Fig. 15:**
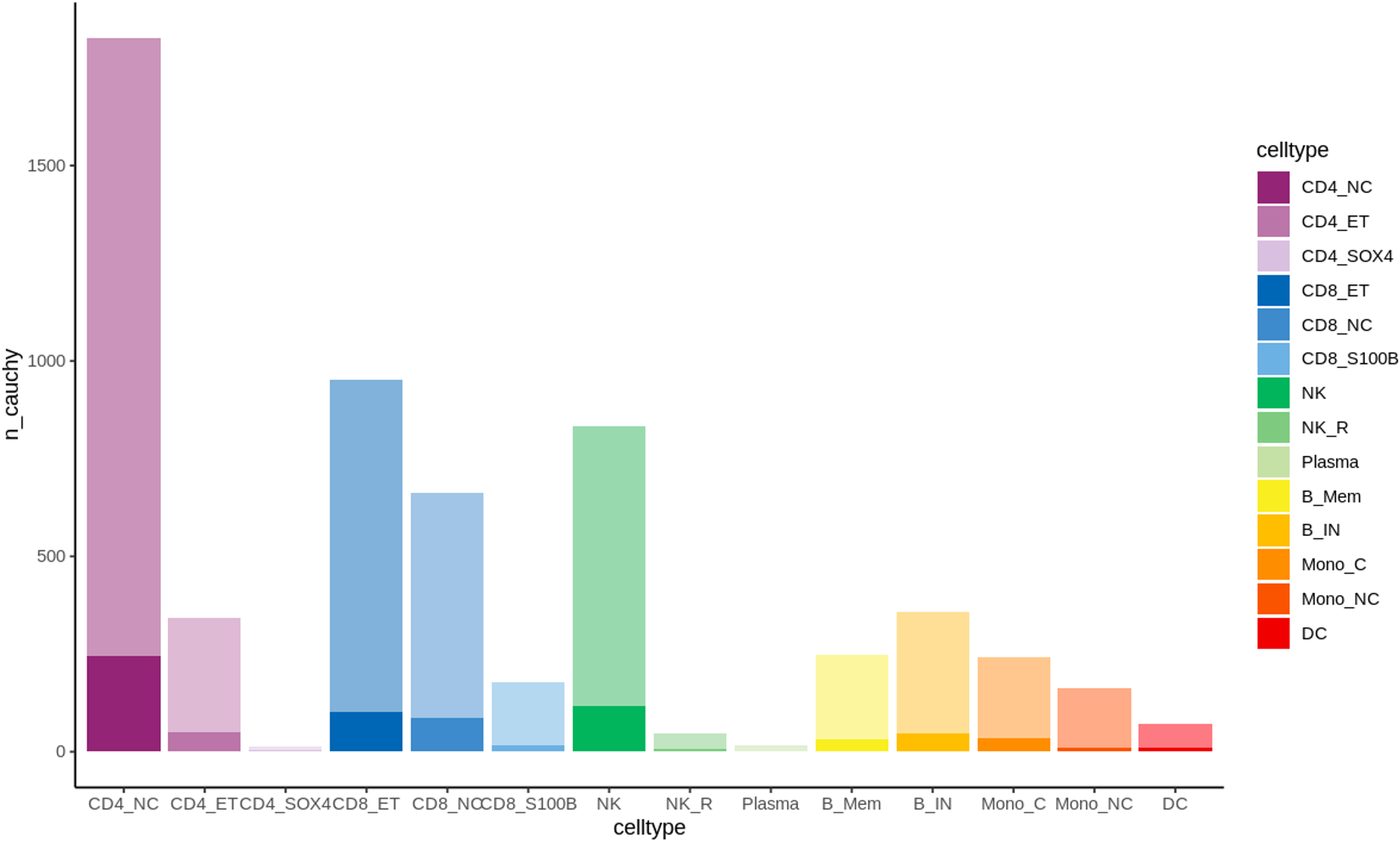
Rare variants conditional analysis results. For each cell type, bars represent the number of eGenes identified by our rare variant analysis (faded colour), and the number of eGenes that remain significant after conditioning on any significant single-variant signal for the same gene (solid colour).

