## Supplementary Note for "Efficient and accurate mixed model association tool for single-cell eQTL analysis"

#### 1 Notations of the SAIGE-QTL method

Assuming the data contain read counts from RNA sequencing mapped to a gene for $N$ cells across $n$ individuals, $c_i$ cells belong to the $ith$ individual and $N = \sum_{i=n}^{n} c_i$. The sequencing read count for the gene of interest, $y_{ij}$, of the $j$th cell from the $i$th individual is assumed to follow Poisson($\mu_{ij}$), where $\mu_{ij} = exp(\eta_{ij})$ and $\eta_{ij} = X_{cij}\alpha_c + X_{di}\alpha_d + G_i\beta + b_i$. Let $G_i$ represent the allele count (0,1, or 2) of the $i$th individual for the genetic variant to test, $X_{cij}$ represent $p_c$ cell-level covariates, such as cell states, including the intercept for the $j$th cell of the $i$th individual, and $X_{di}$ represent $p_d$ individual (donor)-level covariates, such as age, gender, and ancestry PCs, for the $i$th individual. The random effects $\boldsymbol{b_d}$ is a length $n$ vector of $b_i$ and is assumed to follow multivariate Normal distribution $b_i \sim N(0, \sum_{k=1}^{K} \tau_k \boldsymbol{\Psi}_k)$, where $\tau_k$ are the variance component parameters and $\boldsymbol{\Psi}_k$ are known $n \times n$ variance-covariance matrices, which can include the identity matrix to account for intra-individual variance of read counts across multiple cells and the genetic relationship matrix to account for sample relatedness, if any.

The read counts $y_{ij}$ are assumed to be independent conditional on the random effect $(X_{cij}, X_{di}, G_i, b_i)$ and follows the Poisson distribution with mean an variance $E(y_{ij}|X_{cij}, X_{di}, G_i, b_i) = \mu_{ij}$ and $Var(y_{ij}|X_{cij}, X_{di}, G_i, b_i) = \phi v(\mu_{ij})$, where $v(\mu_{ij}) = \mu_{ij}$ is the variance function and the dispersion parameter $\phi = 1$.

Let $\boldsymbol{Z}$ be the $N \times n$ design matrix. Each row in $\boldsymbol{Z}$ contains a 1 and all other elements are 0s to indicate which individual has the cell. The Poisson mixed model can be written as

$$\boldsymbol{\eta} = \boldsymbol{X_c}\boldsymbol{\alpha_c} + \boldsymbol{Z}\boldsymbol{X_d}\boldsymbol{\alpha_d} + \boldsymbol{Z}\boldsymbol{G}\beta + \boldsymbol{b}$$

, where $\boldsymbol{\eta}$ is a length of $N$ vector of $\eta_{ij}$, $\boldsymbol{X_c}$ is a $N \times p_c + 1$ matrix containing cell-level covariates including the intercept, $\boldsymbol{X_d}$ is a $n \times p_d$ matrix containing individual-level covariates, and the random effects for all cells $\boldsymbol{b} = \boldsymbol{Z}\boldsymbol{b_d}$ is a vector with length of $N$ . $\boldsymbol{b} \sim N(0, \sum_{k=1}^{K} \tau_k Z \Psi_k Z^T)$ accounts for all sources of correlation in the data.

##### 1.1 Poisson generalized linear mixed model and penalized quasi-likelihood

Similar algorithms that were used in previous GWAS methods [1, 2, 3] are used in SAIGE-QTL to fit the Poisson generalized linear mixed model under the null hypothesis $H_0 : \beta = 0$. To estimate

$(\boldsymbol{\alpha}, \phi, \boldsymbol{\tau})$, where $\boldsymbol{\alpha} = (\boldsymbol{\alpha_c}, \boldsymbol{\alpha_d})$, the integrated quasi-likelihood function can be written as

$$
\begin{aligned}
L\left(\boldsymbol{\alpha}, \beta=0, \phi, \boldsymbol{\tau}\right) = & \int exp\{\sum_{i=1}^{n}\sum_{j=1}^{c_i} ql_{ij}(\boldsymbol{\alpha}, \beta=0|\boldsymbol{b})\} \\
& \times (2\pi)^{-\frac{N}{2}} \frac{1}{\left|\sum_{k=1}^{K} \tau_k \boldsymbol{V_K}\right|^{1/2}} \exp\left(-\frac{1}{2}\mathbf{b}^\top (\sum_{k=1}^{K} \tau_k \boldsymbol{V_K})^{-1}\mathbf{b}\right) d\mathbf{b} \\
= & (2\pi)^{-\frac{N}{2}} \frac{1}{\left|\sum_{k=1}^{K} \tau_k \boldsymbol{V_K}\right|^{1/2}} \\
& \times \int exp\left(\sum_{i=1}^{n}\sum_{j=1}^{c_i} ql_{ij}(\boldsymbol{\alpha}, \beta=0|\boldsymbol{b}) - \left(\frac{1}{2}\mathbf{b}^\top (\sum_{k=1}^{K} \tau_k \boldsymbol{V_K})^{-1}\mathbf{b}\right)\right) d\mathbf{b}
\end{aligned}
$$

, where $\mathbf{V_k} = \mathbf{Z\Psi_k Z^T}$

$$
ql_{ij}(\boldsymbol{\alpha}, \beta=0|\boldsymbol{b})\} = \int_{y_{ij}}^{\mu_{ij}} \frac{(y_{ij}-\mu)}{\phi v(\mu)} d(\mu) = \int_{y_{ij}}^{\mu_{ij}} \frac{y_{ij}-\mu}{\mu} d(\mu)
$$

is the quasi-likelihood for the $j$th cell from the $i$th individual given the random effect $\boldsymbol{b}$.

Let $f(\boldsymbol{b}) = \sum_{i=1}^{n}\sum_{j=1}^{c_i} ql_{ij}(\boldsymbol{\alpha}, \beta=0|\boldsymbol{b}) - \frac{1}{2}\mathbf{b}^\top (\sum_{k=1}^{K} \tau_k \boldsymbol{V_K})^{-1}\mathbf{b}$, we can then approximate the integral using the Laplace approximation,

$$
\int \exp f(\mathbf{b}) db \approx (2\pi)^{N/2} \left|-f''(\tilde{\mathbf{b}})\right|^{-1/2} \exp(f(\tilde{\mathbf{b}})),
$$

, where $\tilde{\mathbf{b}} = \arg_b \max f(\mathbf{b})$ is the solution to $f'(\mathbf{b}) = 0$. Therefore, the log integrated quasi-likelihood function can be approximated by

$f'(\mathbf{b}) = \frac{\partial \sum_{i=1}^{n}\sum_{j=1}^{c_i} ql_{ij}(\boldsymbol{\alpha},\beta=0|\boldsymbol{b})}{\partial b} = \sum_{i=1}^{n}\sum_{j=1}^{c_i} \frac{\partial ql_{ij}(\boldsymbol{\alpha},\beta=0|\boldsymbol{b})}{\partial b}$

$\partial^2 ql_{ij}(\boldsymbol{\alpha}, \beta=0|\boldsymbol{b})\frac{}{\partial b \partial b^T = -\frac{\partial \mu_{ij}}{\partial \eta_{ij}}\frac{\partial \eta_{ij}}{\partial b_{ij}} Z^T = -\mu_{ij} Z_{ij} Z_{ij}^T}$

$f''(\tilde{\mathbf{b}}) = -(\sum_{i=1}^{n}\sum_{j=1}^{c_i} \mu_{ij} Z_{ij} Z_{ij}^T) - (\sum_{k=1}^{K} \tau_k \boldsymbol{V_K})^{-1}$

$$
\begin{aligned}
ql\left(\alpha, \beta=0, \phi, \boldsymbol{\tau}\right) \approx & -\frac{1}{2}\log\left|\sum_{k=1}^{K} \tau_k \boldsymbol{V_K}\right| - \frac{1}{2}\log\left|(\sum_{i=1}^{n}\sum_{j=1}^{c_i} \mu_{ij} Z_{ij} Z_{ij}^T) + (\sum_{k=1}^{K} \tau_k \boldsymbol{V_K})^{-1}\right| \\
& + \sum_{i=1}^{n}\sum_{j=1}^{c_i} ql_{ij}(\boldsymbol{\alpha}, \beta=0|\tilde{\mathbf{b}}) - \frac{1}{2}\tilde{\mathbf{b}}^\top (\sum_{k=1}^{K} \tau_k \boldsymbol{V_K})^{-1}\tilde{\mathbf{b}} \\
= & -\frac{1}{2}\log\left|\sum_{k=1}^{K} Z(\tau_k \Psi_k \boldsymbol{W} + I)Z^T\right| + \sum_{i=1}^{n}\sum_{j=1}^{c_i} ql_{ij}(\boldsymbol{\alpha}, \beta=0|\tilde{\mathbf{b}}) - \frac{1}{2}\tilde{\mathbf{b}}^\top (\sum_{k=1}^{K} \tau_k \boldsymbol{V_K})^{-1}\tilde{\mathbf{b}}
\end{aligned} \tag{1}
$$

, where $\mathbf{W} = diag(\mu)$

##### 1.2 Estimation of the fixed effects and random effects given the variance components

We can obtain the score functions by taking derivatives of $ql\left(\alpha, \beta=0, \phi, \boldsymbol{\tau}\right)$ with respect to $\beta, \alpha$, and b.

$$\begin{aligned}
\frac{\partial ql\left(\boldsymbol{\alpha}, \beta=0, \phi, \boldsymbol{\tau}\right)}{\partial \boldsymbol{\alpha}} =& \boldsymbol{X}^T\left(\boldsymbol{y}-\boldsymbol{\mu}\right) \\
\frac{\partial ql\left(\boldsymbol{\alpha}, \beta=0, \phi, \boldsymbol{\tau}\right)}{\partial \beta} =& \boldsymbol{G}^T\left(\boldsymbol{y}-\boldsymbol{\mu}\right) \\
\frac{\partial ql\left(\boldsymbol{\alpha}, \beta=0, \phi, \boldsymbol{\tau}\right)}{\partial \boldsymbol{b}} =& (\boldsymbol{y}-\boldsymbol{\mu}) - (\sum_{k=1}^{K} \tau_k \boldsymbol{V_K})^{-1}\boldsymbol{b}
\end{aligned}$$

, where $\boldsymbol{X} = \begin{bmatrix}\boldsymbol{X_c} & \boldsymbol{ZX_d}\end{bmatrix}$. For the score test of $H_0 : \beta = 0$ vs $H_1 : \beta \neq 0$, we only estimate the $(\hat{\alpha}, \hat{\beta}, \hat{\mathbf{b}})$ that maximize $ql\left(\boldsymbol{\alpha}, \beta=0, \phi, \boldsymbol{\tau}\right)$ under $H_0$, setting $\beta = 0$. We denote the working outcome vector $\mathbf{Y} = \boldsymbol{\eta} + \boldsymbol{W}^{-1}(\boldsymbol{y}-\boldsymbol{\mu})$, where $\boldsymbol{W} = diag(\mu_i)$, $\boldsymbol{\eta} = (\eta_1, \ldots, \eta_N)$, $\boldsymbol{\delta} = (\delta_1, \ldots, \delta_N)$, and $\boldsymbol{\mu} = (\mu_1, \ldots, \mu_N)$. Then, $\boldsymbol{y}-\boldsymbol{\mu} = \boldsymbol{W}(\boldsymbol{Y}-\boldsymbol{\eta}) = \boldsymbol{W}(\boldsymbol{Y} - \boldsymbol{X\beta} - \boldsymbol{Zb})$, and the score equations can be written as,

$$\begin{bmatrix}\mathbf{X}^\top\mathbf{W}\mathbf{X} & \mathbf{X}^\top\mathbf{W} \\ \mathbf{W}\mathbf{X} & \mathbf{W} + (\sum_{\mathbf{k=1}}^{\mathbf{K}} \tau_{\mathbf{k}} \boldsymbol{V_K})^{-\mathbf{1}}\end{bmatrix}\begin{bmatrix}\alpha \\ b\end{bmatrix} = \begin{bmatrix}\mathbf{X}^\top\mathbf{W}\mathbf{Y} \\ \mathbf{W}\mathbf{Y}\end{bmatrix}$$

Let $\boldsymbol{\Sigma} = \mathbf{W}^{-\mathbf{1}} + \sum_{\mathbf{k=1}}^{\mathbf{K}} \tau_{\mathbf{k}} \boldsymbol{V_K}$, then

$$\begin{aligned}
\hat{\boldsymbol{\alpha}} &= \left(\mathbf{X}^\top\boldsymbol{\Sigma}^{-1}\mathbf{X}\right)^{-\mathbf{1}}\mathbf{X}^\top\boldsymbol{\Sigma}^{-1}\mathbf{Y} \\
\hat{\mathbf{b}} &= \left(\sum_{\mathbf{k=1}}^{\mathbf{K}} \tau_{\mathbf{k}} \boldsymbol{V_K}\right)\boldsymbol{\Sigma}^{-\mathbf{1}}\left(\mathbf{Y} - \mathbf{X}\hat{\boldsymbol{\alpha}}\right).
\end{aligned}$$

##### 1.3 Estimation of the variance component

Given $\hat{\boldsymbol{\alpha}} = \hat{\boldsymbol{\alpha}}(\boldsymbol{\tau}), \hat{\boldsymbol{b}} = \hat{\boldsymbol{b}}(\boldsymbol{\tau})$ estimated, from (1) the log-likelihood of the variance component can be derived as,

$$ql(\hat{\boldsymbol{\alpha}}(\boldsymbol{\tau}, \phi), \beta=0, \phi, \boldsymbol{\tau}) = c - \frac{1}{2}log|\boldsymbol{\Sigma}| - \frac{\mathbf{1}}{\mathbf{2}}\mathbf{Y}^\top\mathbf{P}\mathbf{Y},$$

where $\mathbf{P} = \boldsymbol{\Sigma}^{-\mathbf{1}} - \boldsymbol{\Sigma}^{-\mathbf{1}}\mathbf{X}\left(\mathbf{X}^\top\boldsymbol{\Sigma}^{-\mathbf{1}}\mathbf{X}\right)^{-\mathbf{1}}\mathbf{X}^\top\boldsymbol{\Sigma}^{-\mathbf{1}}$ and $\boldsymbol{\Sigma} = \boldsymbol{W}^{-\mathbf{1}} + \sum_{\boldsymbol{k=1}}^{\boldsymbol{K}} \boldsymbol{\tau_k V_k}$. We maximize the corresponding restricted maximum-likelihood (REML),

$$ql_R(\hat{\boldsymbol{\alpha}}(\boldsymbol{\tau}, \phi), \beta=0, \phi, \boldsymbol{\tau}) = c_R - \frac{1}{2}log|\boldsymbol{\Sigma}| - \frac{1}{2}log|\mathbf{X}^\top\boldsymbol{\Sigma}^{-\mathbf{1}}\mathbf{X}| - \frac{\mathbf{1}}{\mathbf{2}}\mathbf{Y}^\top\mathbf{P}\mathbf{Y}.$$

The score function with respect to $\tau$ are given by,

$$\mathbf{U}_\tau = \frac{\partial ql_R(\hat{\boldsymbol{\alpha}}(\boldsymbol{\tau}, \phi), \beta=0, \phi, \boldsymbol{\tau})}{\partial \tau_k} = \frac{1}{2}\left[\mathbf{Y}^\top\mathbf{PZ}\boldsymbol{\Psi}_{\mathbf{k}}\mathbf{Z}^{\mathbf{T}}\mathbf{PY} - tr(\mathbf{PZ}\boldsymbol{\Psi}_{\mathbf{k}}\mathbf{Z}^{\mathbf{T}})\right].$$

The corresponding observed information function, and the expected information function are given by

$$\begin{aligned}
\mathbf{J}_\tau &= -\frac{\partial^2 ql_R(\hat{\boldsymbol{\alpha}}(\boldsymbol{\tau}, \phi), \beta=0, \phi, \boldsymbol{\tau})}{\partial \tau_k^2} = -\frac{1}{2}tr(\mathbf{PV_kPV_k}) + \mathbf{Y}^\top\mathbf{PV_kPV_kPY}, \\
E\left(\mathbf{J}_\tau\right) &= E\left[-\frac{\partial^2 ql_R(\hat{\boldsymbol{\alpha}}(\boldsymbol{\tau}, \phi), \beta=0, \phi, \boldsymbol{\tau})}{\partial \tau_k^2}\right] = \frac{1}{2}tr(\mathbf{PV_kPV_k}),
\end{aligned}$$

respectively. Evaluating both observed and expected information functions involves computationally expensive trace computations. To avoid the trace computations, the average information is used in the AI-REML[1, 2, 4] algorithm. The average information is expressed as the average of $J_\tau$ and $E(J_\tau)$,

$$\mathbf{AI}_{\tau_k\tau_q} = \frac{1}{2}\mathbf{Y}^\top\mathbf{PV_kPV_qPY}.$$

, where $\mathbf{AI}$ is an $K \times K$ matrix with the $(k, q)$-th element $\mathbf{AI}_{\tau_k\tau_q}$.

##### 1.3.1 Algorithm to fit the null mixed model

The null model fitting algorithm can be summarized as,

1. Fit a Poisson linear model with $\boldsymbol{\tau} = 0$ to get initial estimates $\hat{\boldsymbol{\alpha}}^{(0)}$ and working outcome vector $Y^{(0)}$.
2. At the $i$-th step, update $\hat{\boldsymbol{\tau}}$ using $\hat{\boldsymbol{\tau}}^{(i)} = \hat{\boldsymbol{\tau}}^{(i-1)} + \left\{ \mathbf{AI}_{\boldsymbol{\tau}}\Big|_{\boldsymbol{\tau}=\hat{\boldsymbol{\tau}}^{(i-1)}} \right\}^{-1} \left\{ \mathbf{U}_{\boldsymbol{\tau}}\Big|_{\boldsymbol{\tau}=\hat{\boldsymbol{\tau}}^{(i-1)}} \right\}$.
3. Update $\hat{\boldsymbol{\alpha}}$, $\hat{\boldsymbol{b}}$ using $\mathbf{Y}$ and $\hat{\boldsymbol{\tau}}^{(i)}$.
4. Update $\mathbf{Y}$ using $\hat{\boldsymbol{b}}^{(i)}, \hat{\boldsymbol{\alpha}}^{(i)}, \hat{\boldsymbol{\tau}}^{(i)}$.
5. Repeat steps 2–4, until $\max\left\{ \frac{|\hat{\beta}^{(i)} - \hat{\beta}^{(i-1)}|}{|\hat{\beta}^{(i)}| + |\hat{\beta}^{(i-1)}|}, \frac{|\hat{\boldsymbol{\tau}}^{(i)} - \hat{\boldsymbol{\tau}}^{(i-1)}|}{|\hat{\boldsymbol{\tau}}^{(i)}| + |\hat{\boldsymbol{\tau}}^{(i-1)}|} \right\} \leq tolerance.$

#### 1.4 Score test

The score test statistic under the null hypothesis is given by,

$$T = \frac{\partial ql}{\partial \beta}\Big|_{(\hat{b}, \beta=0, \hat{\alpha}, \hat{\tau})} = \mathbf{G}^{\top}\mathbf{Z}^{\top}(\mathbf{y} - \hat{\mu}) = \tilde{\mathbf{G}}^{\top}(\mathbf{y} - \hat{\mu}), \tag{2}$$

where $\tilde{\mathbf{G}} = \mathbf{ZG} - \mathbf{X}\left(\mathbf{X}^{\top}\hat{\mathbf{W}}\mathbf{X}\right)^{-\mathbf{1}}\mathbf{X}^{\top}\hat{\mathbf{W}}\mathbf{ZG}$ is the covariate adjusted genotype vector, and $\boldsymbol{X} = \begin{bmatrix}\boldsymbol{X_c} & \boldsymbol{ZX_d}\end{bmatrix}$ is the covariate matrix with intercept included. The information matrix corresponding to the score equations in (2) is given by,

$$\mathbf{I}(\alpha, \beta, b) = \begin{bmatrix} \mathbf{X}^{\top}\mathbf{WX} & \mathbf{X}^{\top}\mathbf{WG} & \mathbf{X}^{\top}\mathbf{W} \\ \mathbf{G}^{\top}\mathbf{WX} & \mathbf{G}^{\top}\mathbf{WG} & \mathbf{G}^{\top}\mathbf{W} \\ \mathbf{WX} & \mathbf{Z}^{\top}\mathbf{WG} & \mathbf{W} + (\sum_{\mathbf{k=1}}^{\mathbf{K}} \tau_{\mathbf{k}}\mathbf{V_k})^{-\mathbf{1}} \end{bmatrix},$$

$A_{ij}$ and $A_{\bullet i}$ denote the $(i,j)$-th element and $i$-th column of a matrix $A$, respectively. Then, the variance of the score statistic under $H_0$ is given by,

$$Var_{H_0}(T) = \left(\mathbf{I}(\hat{\alpha}, \hat{\beta}, \hat{\mathbf{b}})^{-\mathbf{1}}\right)_{\mathbf{22}} = \mathbf{G}^{\top}\hat{\mathbf{P}}\mathbf{G} = \tilde{\mathbf{G}}^{\top}\hat{\mathbf{P}}\tilde{\mathbf{G}},$$

where $\hat{\mathbf{P}} = \hat{\mathbf{\Sigma}}^{-\mathbf{1}} - \hat{\mathbf{\Sigma}}^{-\mathbf{1}}\mathbf{X}\left(\mathbf{X}^{\top}\mathbf{\Sigma}^{-\mathbf{1}}\mathbf{X}\right)^{-\mathbf{1}}\mathbf{X}^{\top}\hat{\mathbf{\Sigma}}^{-\mathbf{1}}$, $\hat{\mathbf{\Sigma}} = \hat{\mathbf{W}}^{-\mathbf{1}} + \sum_{\mathbf{k=1}}^{\mathbf{K}} \hat{\tau}_{\mathbf{k}}\mathbf{V_k}$, and $\mathbf{V_k} = \mathbf{Z\Psi_k Z^T}$.

### 2 Variance ratio approximation

Computation of the variance of the score statistic $Var_{H_0}(T) = \tilde{\mathbf{G}}^{\top}\hat{\mathbf{P}}\tilde{\mathbf{G}}$ requires calculating $\hat{\mathbf{P}}\tilde{\mathbf{G}}$ repeatedly for all markers, which is computationally expensive. To avoid calculating $\hat{\mathbf{P}}\tilde{\mathbf{G}}$ for all the markers, previous linear mixed model [5, 6, 7] and logistic mixed model methods [2] first estimate the variance ratio $\hat{r} = \tilde{\mathbf{G}}^{\top}\hat{\mathbf{P}}\tilde{\mathbf{G}}/\tilde{\mathbf{G}}^{\top}\hat{\mathbf{W}}\tilde{\mathbf{G}}$ using a small set of markers, and then approximate the variance of the score statistic for all markers by $\hat{r}\tilde{\mathbf{G}}^{\top}\hat{\mathbf{W}}\tilde{\mathbf{G}}$ in step 2. It has been shown in the studies that $\hat{r}$ is approximately constant for all genetic variants. This saves substantial computation time since $\hat{\mathbf{W}}$ is a diagonal matrix. Following the Computation of the variance of the score statistic $Var_{H_0}(T) = \tilde{\mathbf{G}}^{\top}\hat{\mathbf{P}}\tilde{\mathbf{G}}$ requires calculating $\hat{\mathbf{P}}\tilde{\mathbf{G}}$ repeatedly for all markers, which is computationally expensive. To avoid calculating $\hat{\mathbf{P}}\tilde{\mathbf{G}}$ for all the markers, we estimate the variance ratio $\hat{r} = \tilde{\mathbf{G}}^{\top}\hat{\mathbf{P}}\tilde{\mathbf{G}}/\tilde{\mathbf{G}}^{\top}\hat{\mathbf{W}}\tilde{\mathbf{G}}$ using a small set of markers, and then approximate the variance of the score statistic for all markers by $\hat{r}\tilde{\mathbf{G}}^{\top}\hat{\mathbf{W}}\tilde{\mathbf{G}}$ in step 2. This saves substantial computation time since $\hat{\mathbf{W}}$ is a diagonal matrix. Such variance ratio approximation approaches were previously used in various linear[5, 6, 7] and logistic mixed effects models[2] to speed up computation. Following the theoretical justification by Dey et al.[3] to show the approximation also works in the frailty models, we can show that it also works in the Poisson mixed model for eQTL mapping with scRNA-seq data.

Let $E(G_i) = \mu_g$ and the covariance matrix of $\mathbf{G}$ is given by $\sigma_g^2 \mathbf{\Psi}$, where $\mathbf{\Psi}$ is the correlation matrix of $\mathbf{G}$ and represents the $n \times n$ kinship matrix. The exact characterization of $\mathbf{\Psi}$ is not needed for the proof. Then the elements of $\mathbf{G}$ follows the $Bin(2, p_g)$ distribution, then $\mu_g = 2p_g$, and $\sigma_g^2 = 2p_g(1-p_g)$. Let $\mathbf{G_c}$ be the $n \times 1$ unadjusted (but mean-centered) genotype vector. Then $E(\mathbf{G_c}) = 0$ and $Cov(\mathbf{G_c}) = \sigma_g^2 \mathbf{\Psi}$. Let $\mathbf{Q_{\tilde{X}}} = \mathbf{X}(\mathbf{X}^\top \hat{\mathbf{W}} \mathbf{X})^{-\mathbf{1}} \mathbf{X}^\top \hat{\mathbf{W}}$ be the weighted projection matrix. Then, $E(\tilde{\mathbf{G}}) = (\mathbf{I} - \mathbf{Q_{\tilde{X}}})\mathbf{Z}\mu_g \mathbf{1} = 0$, and $Cov(\tilde{\mathbf{G}}) = \sigma_g^2 (\mathbf{I} - \mathbf{Q_{\tilde{X}}})\mathbf{Z}\mathbf{\Psi}\mathbf{Z}^\mathbf{T}(\mathbf{I} - \mathbf{Q_{\tilde{X}}})^\top$, where $\mathbf{1}$ is the $n \times 1$ vector of all element equal to unity. We scale both the numerator and denominator of the variance ratio by $N^{-1}$ so that they don't blow to infinity when looked at individually. Then, for the numerator,

$$E(N^{-1}\tilde{\mathbf{G}}^\top \hat{\mathbf{P}} \tilde{\mathbf{G}}) = \frac{\sigma_g^2}{N} tr\left[\hat{\mathbf{P}}(\mathbf{I} - \mathbf{Q_{\tilde{X}}})\mathbf{Z}\mathbf{\Psi}\mathbf{Z}^\mathbf{T}(\mathbf{I} - \mathbf{Q_{\tilde{X}}})^\top\right] = \frac{\sigma_g^2}{N} tr(\hat{\mathbf{P}}\mathbf{Z}\mathbf{\Psi}\mathbf{Z}^\mathbf{T}),$$

since $(\mathbf{I} - \mathbf{Q_{\tilde{X}}})^\top \hat{\mathbf{P}} (\mathbf{I} - \mathbf{Q_{\tilde{X}}}) = \hat{\mathbf{P}}$. Similarly, for the denominator,

$$E(N^{-1}\mathbf{G_c}^\top \mathbf{Z}^\top \hat{\mathbf{W}} \mathbf{Z} \mathbf{G_c}) = \frac{\sigma_g^2}{N} tr\Big(\mathbf{Z}^\top \hat{\mathbf{W}} \mathbf{Z} \mathbf{\Psi}\Big),$$

As the eigenvalues of $\hat{\mathbf{P}}, \hat{\mathbf{W}}, \mathbf{\Psi}$ are bounded, and the distribution of $\mathbf{G}$ has bounded support, the variances of the numerator and the denominator terms are both $O(N^{-1})$, and the variance ratio converges to,

$$\hat{r} = \frac{\tilde{\mathbf{G}}^\top \hat{\mathbf{P}} \tilde{\mathbf{G}}}{\mathbf{G_c}^\top \mathbf{Z}^\top \hat{\mathbf{W}} \mathbf{Z} \mathbf{G_c}} \xrightarrow{p} \frac{lim_{N\to\infty}\left\{N^{-1} tr(\hat{\mathbf{P}}\mathbf{Z}\mathbf{\Psi}\mathbf{Z}^\mathbf{T})\right\}}{lim_{N\to\infty}\left\{N^{-1} tr\Big(\mathbf{Z}^\top \hat{\mathbf{W}} \mathbf{Z} \mathbf{\Psi}\Big)\right\}}.$$

The ratio on the right-hand side is constant across all markers as the individual limits in the numerator and denominator exist and are bounded away from zero.

In addition, as we have previously shown in SAIGE-GENE[8], the variation of the estimated variance ratio can be smaller when $\hat{\mathbf{\Sigma}}^{-\mathbf{1}}$ is used in the denominator than when only $\hat{\mathbf{W}}$ is used. Therefore, in SAIGE-QTL, we incorporate $\hat{\mathbf{\Sigma}}^{-\mathbf{1}}$ and the covariate adjusted genotype vector into the denominator to approximate the $Var_{H_0}(T)$ with a smaller variation. More specifically, the variance ratio $\hat{r_s} = \tilde{\mathbf{G}}^\top \hat{\mathbf{P}} \tilde{\mathbf{G}} / \tilde{\mathbf{G}}_\mathbf{d}^\top \hat{\mathbf{\Sigma}}^{-\mathbf{1}} \tilde{\mathbf{G}}_\mathbf{d}$ is estimated. Let $\boldsymbol{X_d} = [\mathbf{1} \quad \boldsymbol{X_d}]$ represent the donor-level covariate matrix with intercept included with the size $n \times (p_d + 1)$, then $\tilde{\mathbf{G}}_\mathbf{d}$ is the donor-level covariate adjusted genotype vector with size $N \times 1$ and

$$\begin{aligned}\tilde{\mathbf{G}}_d &= \mathbf{Z}\mathbf{G} - \mathbf{Z}\tilde{\mathbf{X}}_\mathbf{d}\left(\tilde{\mathbf{X}}_\mathbf{d}^\top \mathbf{Z}^\top \hat{\mathbf{W}} \mathbf{Z} \tilde{\mathbf{X}}_\mathbf{d}\right)^{-\mathbf{1}} \tilde{\mathbf{X}}_\mathbf{d}^\top \mathbf{Z}^\top \hat{\mathbf{W}} \mathbf{Z} \mathbf{G} \\ &= \mathbf{Z}(\mathbf{G} - \tilde{\mathbf{X}}_\mathbf{d}\left(\tilde{\mathbf{X}}_\mathbf{d}^\top \mathbf{Z}^\top \hat{\mathbf{W}} \mathbf{Z} \tilde{\mathbf{X}}_\mathbf{d}\right)^{-\mathbf{1}} \tilde{\mathbf{X}}_\mathbf{d}^\top \mathbf{Z}^\top \hat{\mathbf{W}} \mathbf{Z} \mathbf{G})\end{aligned}$$

. Similarly, Let $\mathbf{Q}_{\tilde{\mathbf{X}}_\mathbf{d}} = \tilde{\mathbf{X}}_\mathbf{d}\left(\tilde{\mathbf{X}}_\mathbf{d}^\top \mathbf{Z}^\top \hat{\mathbf{W}} \mathbf{Z} \tilde{\mathbf{X}}_\mathbf{d}\right)^{-\mathbf{1}} \tilde{\mathbf{X}}_\mathbf{d}^\top \mathbf{Z}^\top \hat{\mathbf{W}} \mathbf{Z}$ be the weighted projection matrix to project out the donor-level covariates with intercept, then $\tilde{\mathbf{G}}_\mathbf{d} = \mathbf{Z}(\mathbf{I} - \mathbf{Q}_{\tilde{\mathbf{X}}_\mathbf{d}})\mathbf{G}$, $E(\tilde{\mathbf{G}}_\mathbf{d}) = \mathbf{Z}(\mathbf{I} - \mathbf{Q}_{\tilde{\mathbf{X}}_\mathbf{d}})\mu_g \mathbf{1} = 0$, and $Cov(\tilde{\mathbf{G}}_\mathbf{d}) = \sigma_g^2 \mathbf{Z}(\mathbf{I} - \mathbf{Q}_{\tilde{\mathbf{X}}_\mathbf{d}})\mathbf{\Psi}(\mathbf{I} - \mathbf{Q}_{\tilde{\mathbf{X}}_\mathbf{d}})^\top \mathbf{Z}^\top$.

For the denominator,

$$E(N^{-1}\tilde{\mathbf{G}}_\mathbf{d}^\top \hat{\mathbf{\Sigma}}^{-\mathbf{1}} \tilde{\mathbf{G}}_\mathbf{d}) = \frac{\sigma_g^2}{N} tr\Big(\hat{\mathbf{\Sigma}}^{-\mathbf{1}} \mathbf{Z}(\mathbf{I} - \mathbf{Q}_{\tilde{\mathbf{X}}_\mathbf{d}})\mathbf{\Psi}(\mathbf{I} - \mathbf{Q}_{\tilde{\mathbf{X}}_\mathbf{d}})^\top \mathbf{Z}^\top\Big),$$

Similarly, as the eigenvalues of $\hat{\mathbf{P}}, \hat{\mathbf{W}}, \mathbf{\Psi}, \hat{\mathbf{\Sigma}}^{-\mathbf{1}}$ are bounded, and the distribution of $\mathbf{G}$ has bounded support, the variances of the numerator and the denominator terms are both $O(N^{-1})$, and the variance ratio converges to,

$$\hat{r}_s = \frac{\tilde{\mathbf{G}}^\top \hat{\mathbf{P}} \tilde{\mathbf{G}}}{\tilde{\mathbf{G}}_\mathbf{d}^\top \hat{\mathbf{\Sigma}}^{-\mathbf{1}} \tilde{\mathbf{G}}_\mathbf{d}} \xrightarrow{p} \frac{lim_{N\to\infty}\left\{N^{-1} tr(\hat{\mathbf{P}}\mathbf{Z}\mathbf{\Psi}\mathbf{Z}^\mathbf{T})\right\}}{lim_{N\to\infty}\left\{N^{-1} tr\Big(\hat{\mathbf{\Sigma}}^{-\mathbf{1}} \mathbf{Z}(\mathbf{I} - \mathbf{Q}_{\tilde{\mathbf{X}}_\mathbf{d}})\mathbf{\Psi}(\mathbf{I} - \mathbf{Q}_{\tilde{\mathbf{X}}_\mathbf{d}})^\top \mathbf{Z}^\top\Big)\right\}}.$$

, which is constant across all markers as the individual limits in the numerator and denominator exist and are bounded away from zero.

For each genetic variant, $\hat{r}$ is first used to approximate the variance $Var_{H_0}(T)$, if the p-value ¡ 0.05, $\hat{r_s}$ is used to obtain a more accurate $Var_{H_0}(T)$.

#### 3 Using saddlepoint approximation[9] (SPA) for the null distribution of the score statistic

In traditional score tests, the distribution of the score statistic under $H_0$ is approximated by a Normal distribution, which uses the first two moments, mean and variance. This approach can perform poorly in the tail regions, especially if the underlying distribution is highly skewed when the studying event is very rare or the testing genetic variant has a very low minor allele count (MAC). Here, similar to what has been applied in the logistic mixed models [10, 11, 2, 12] previously, we use the SPA to approximate the null distribution of the score statistic to obtain accurate p-values. For this, we utilize the fact that given the random effects $\boldsymbol{b}$, the phenotype Y, which are read counts from RNA sequencing, independently follow Poisson distribution. $T_{adj} = \tilde{\mathbf{G}}^\top(\mathbf{y} - \hat{\mu})/\sqrt{\hat{r}\tilde{\mathbf{G}}^\top\hat{\mathbf{W}}\tilde{\mathbf{G}}}$ is a weighted sum of independent Poisson random variable. The approximated cumulant generating function (CGF) of $T_{adj}$ is

$$K(\xi;\hat{\mu},c) = \sum_{i=1}^{N}\hat{\mu}_i(e^{\tilde{G}_i c\xi} - \tilde{G}_i c\xi - 1)$$

, where $c = (\tilde{G}^\top W\tilde{G})^{-1/2}$.$K'(\xi;\hat{\mu},c)$ and $K''(\xi;\hat{\mu},c)$ are first and second derivatives of $K$ with( respect to $\xi$.

$$K'(\xi;\hat{\mu},c) = \sum_{i=1}^{N}\hat{\mu}_i\tilde{G}_i c(e^{\tilde{G}_i c\xi} - 1),$$

$$K''(\xi;\hat{\mu},c) = \sum_{i=1}^{N}\hat{\mu}_i\tilde{G}_i^2 c^2 e^{\tilde{G}_i c\xi}$$

To calculate the probability that $T_{adj} < s$, where $s$ is the observed test statistic, we use the following formula

$$Pr(T < s) = \Phi\left\{w + \frac{1}{w}log\left(\frac{v}{w}\right)\right\},$$

where $w = sign(\hat{\xi})[2\{\hat{\xi}s - K(\hat{\xi})\}]^{\frac{1}{2}}$, $v = \hat{\xi}[K''(\hat{\xi})]^{\frac{1}{2}}$, $\hat{\xi}$ is the solution of $K'(\hat{\xi}) = s$, and $\Phi$ is the standard normal distribution function.

#### 4 Approaches to reduce computation and memory cost

We developed SAIGE-QTL based on the SAIGE framework, which utilizes several state-of-the-art approaches to reduce computation and memory cost to fit the null GLMM and to test for genetic associations. Given that in the single-cell RNA sequencing data, each individual may have hundreds and thousands of cells ($N >> n$), which $N$ is the total number of cells and $n$ is the total number of individuals, we use several approaches to make SAIGE-QTL conduct matrix operations on the scale of number of individuals to reduce a computation cost from $O(N)$ to $O(n)$.

First, to compute the test statistics $\mathbf{T} = \mathbf{G}^\top\mathbf{Z}^\top(\mathbf{y} - \hat{\mu})$ for each genetic variant, we pre-compute the statistics $\mathbf{Z}^\top(\mathbf{y} - \hat{\mu})$, so the computation cost of obtaining $\mathbf{T}$ is $O(n)$.

Second, to calculate $Var_{H_0}(T) = \tilde{\mathbf{G}}^\top\hat{\mathbf{P}}\tilde{\mathbf{G}}$ when estimating the variance ratios using randomly selected genetic markers, we need to compute quantities of the form $\hat{\boldsymbol{\Sigma}}^{-1}\mathbf{a}$, where $\mathbf{a}$ is a vector of length $N$. The standard computation technique of inverting $\hat{\boldsymbol{\Sigma}}$ (computation cost $O(N^3)$) and multiplying $\hat{\boldsymbol{\Sigma}}^{-1}$ with $\mathbf{a}$ can be extremely time consuming when $N$ is large. To compute quantities of the form $\hat{\boldsymbol{\Sigma}}^{-1}\mathbf{a}$, we implemented the pre-conditioned conjugate gradient[13] (PCG) method, which computes $\hat{\boldsymbol{\Sigma}}^{-1}\mathbf{a} = \mathbf{x}$ by iteratively solving the linear system of equations $\hat{\boldsymbol{\Sigma}}\mathbf{x} = \mathbf{a}$. Only $\hat{\boldsymbol{\Sigma}}\mathbf{a}$ needs to be computed when the PCG method is used. Instead of storing the $N \times N$ variance-covariance matrix $\mathbf{V}$(memory cost $O(N^2)$), we only store the $K$ matrices $\boldsymbol{\Psi}_\mathbf{k}$ with the size $n \times n$ (memory cost $O(kn^2)$) and the $N \times n$ design matrix $\mathbf{Z}$, which only cost $4N$ bytes if stored using a sparse matrix with 1s. Then, $\hat{\boldsymbol{\Sigma}}\mathbf{a}$ involves the computation for $\mathbf{Z}\boldsymbol{\Psi}_\mathbf{k}\mathbf{Z}^\top\mathbf{a}$, which is computed consecutively, with the time cost O(Nnk). As implemented in SAIGE, we use the Hutchinson's randomized trace estimation[14, 15] for calculating $tr(\mathbf{PV})$.

We also implemented multithreaded parallel computation for the matrix-vector multiplications in the PCG steps using Intel Threading Building Block (TBB) from the RcppParallel[16] package.

Third, as described in , in step 2, the variance of the score statistic for all markers can be approximated by $\hat{r}\mathbf{G_c}^\top\mathbf{Z}^\top\hat{\mathbf{W}}\mathbf{Z}\mathbf{G_c}$ or $\hat{r}_s\tilde{\mathbf{G}}_\mathbf{d}^\top\hat{\mathbf{\Sigma}}^{-1}\tilde{\mathbf{G}}_\mathbf{d}$. We pre-compute both $\mathbf{Z}^\top\hat{\mathbf{W}}\mathbf{Z}$ and $\mathbf{Z}^\top\hat{\mathbf{\Sigma}}^{-1}\mathbf{Z}$, so the computation to approximate the variance of the score statistic becomes $O(n)$ and for $\sim 5\%$ genetic variants, it costs $O(n(p_d + 1))$.

Fourth, to reduce the redundancies of reading genotypes for each genetic variant when mapping eQTLs for all 20,000 genes, SAIGE-QTL allows for analyzing multiple genes. We observed that computation time has been dropped dramatically compared to analyzing each gene separately. This is particukary useful for conducting genome-wide trans-eQTL mapping.

[16] JJ Allaire, Romain Francois, Kevin Ushey, Gregory Vandenbrouck, Marcus Geelnard, and Intel. *RcppParallel: Parallel Programming Tools for 'Rcpp'*, 2019. R package version 4.4.4.
